# Post-acute COVID-19 sequelae in cases managed in the community or hospital in the UK: a population based study

**DOI:** 10.1101/2021.04.09.21255199

**Authors:** Hannah R Whittaker, Claudia Gulea, Ardita Koteci, Constantinos Kallis, Ann D Morgan, Chukwuma Iwundu, Mark Weeks, Rikisha Gupta, Jennifer K Quint

## Abstract

**Objective:** To compare post-COVID-19 sequelae between hospitalised and non-hospitalised individuals

**Design:** Population-based cohort study

**Setting:** 1,383 general practices in England contributing to Clinical Practice Research Database Aurum

**Participants:** 46,687 COVID-19 cases diagnosed between 1^st^ August to 17^th^ October 2020 (45.4% male; mean age 40), either hospitalised within two weeks of diagnosis or non-hospitalised, and followed-up for a maximum of three months.

**Main outcome measures:** Event rates of new symptoms, diseases, prescriptions and healthcare utilisation in hospitalised and non-hospitalised individuals, with between-group comparison using Cox regression. Outcomes compared at 6 and 12 months prior to index date, equating to first UK wave and pre-pandemic. Non-hospitalised group outcomes stratified by age and sex.

**Results:** 45,272 of 46,687 people were non-hospitalised; 1,415 hospitalised. Hospitalised patients had higher rates of 13/26 symptoms and 11/19 diseases post-COVID-19 than the community group, received more prescriptions and utilised more healthcare. The largest differences were noted for rates per 100,000 person-weeks [95%CI] of *breathlessness:* 536 [432 to 663] v. 85 [77 to 93]; *joint pain:* 295 [221 to 392] v. 168 [158 to 179]; *diabetes:* 303 [225 to 416] v. 36 [32 to 42], *hypertension:* 244 [178 to 344] v. 47 [41 to 53]. Although low, rates of chest tightness, tinnitus and lung fibrosis were higher in the community group. 4.2% (1882/45,272) of the community group had a post-acute burden, most frequently reporting anxiety, breathlessness, chest pain and fatigue. In those non-hospitalised, age and sex differences existed in outcome rates. Healthcare utilisation in the community group increased 28.5% post-COVID-19 relative to pre-pandemic.

**Conclusions:** Post-COVID-19 sequelae differ between hospitalised and non-hospitalised individuals, with age and sex-specific differences in those non-hospitalised. Most COVID-19 cases managed in the community do not report ongoing issues to healthcare professionals. Post-COVID-19 follow-up and management strategies need to be tailored to specific groups.

## Introduction

The COVID-19 pandemic continues to cause a pressing and ongoing challenge to global public health. While over 100 million have recovered worldwide, it is increasingly acknowledged the effects of COVID-19 can ripple beyond the acute presentation, with growing evidence of chronic symptoms and new multisystem disease having implications for future health service planning.^1^ Termed ‘long Covid’ or ‘post-acute Covid syndrome’, various prevalence estimates are reported. The Office for National Statistics estimates that over a four week period, 1.1 million individuals in the UK self-reported long COVID-19 symptoms persisting over four weeks, with 13.7% of 20 000 individuals continuing to experience symptoms >12 weeks.^2^ Self-reported symptom data from the Covid Symptom Study smartphone app of over 4000 patients mainly across the UK and US noted lower estimates.^3^ In addition to persistent symptoms, evidence is also emerging of new end-organ dysfunction in those who recover from acute infection, with a potential to negatively impact cardiovascular,^4^ respiratory,^5^ metabolic,^6^ haematological,^7^ psychological,^8^ and neurological health.^9^

While working definitions of long Covid have been established, such as recently by NICE (National Institute for Clinical Excellence),^10^ understanding of short and long-term health consequences post-COVID-19 infection remains limited. One of the main factors precluding a comprehensive understanding of post-Covid-sequelae is the predominant focus in published literature and ongoing longitudinal studies on assessing outcomes in hospitalised patients.^11^ Yet, it is estimated that 80% of COVID-19 cases are mild, with only 3.5% of cases in England requiring hospitalisation at the start of the first wave,^12^ and 10.5% requiring hospitalisation cumulatively since the start of pandemic.^13^ Notably, patient group letters,^14^ surveys,^15^ and qualitative studies highlight a high proportion of patients with persistent debilitating symptoms who either had no access to testing or tested either positive or negative yet were not hospitalised.^16^ Few studies have compared outcomes across the spectrum of COVID-19 severity post-acute infection, with findings to date limited in generalisability owing to small cohort sizes and selection biases.^15^

Understanding the nature and burden of post-COVID-19 sequelae across different patient groups is crucial to shaping effective rehabilitation services that can provide adequate and tailored support to those affected. In this study, we used a large UK primary care longitudinal dataset, broadly representative of the UK population, to investigate new primary care-recorded symptoms, diseases, prescriptions and healthcare utilisation in patients post-acute COVID-19 infection, comparing outcomes between those managed in the community and those hospitalised.

## Methods

### Data Source

We used the Clinical Practice Research Database (CPRD) Aurum, a nationally representative database of anonymised primary care electronic healthcare records, which holds data on symptoms, diagnoses, prescriptions, test results, immunisations, consultations, hospitalisations and specialist referrals for over 39 million patients across the UK, covering approximately 19% of the UK population.^17^ It is one of the largest longitudinal databases worldwide and its use has been extensively validated.^18^ Clinical information is entered using SNOMED CT codes, while prescriptions are recorded using British National Formulary codes. Data from patient records are only used if of a certain standard of quality. For participating general practices, data from secondary care encounters is also fed back into primary care records and CPRD Aurum.

### Study Population

The study population included individuals aged 18 years or over, registered with a general practice contributing to CPRD Aurum. Cases of COVID-19 were identified from 1st August - 17th October 2020.

Patients’ index date was their COVID-19 diagnosis date. Eligible patients were categorised as community or hospitalised COVID-19 patients, depending on hospital admission for COVID-19 within two weeks of index date. Patients were followed up to three months, with censoring at the earliest of transfer out of practice, death or end of follow-up (Fig. 1).

**Fig. 1:**
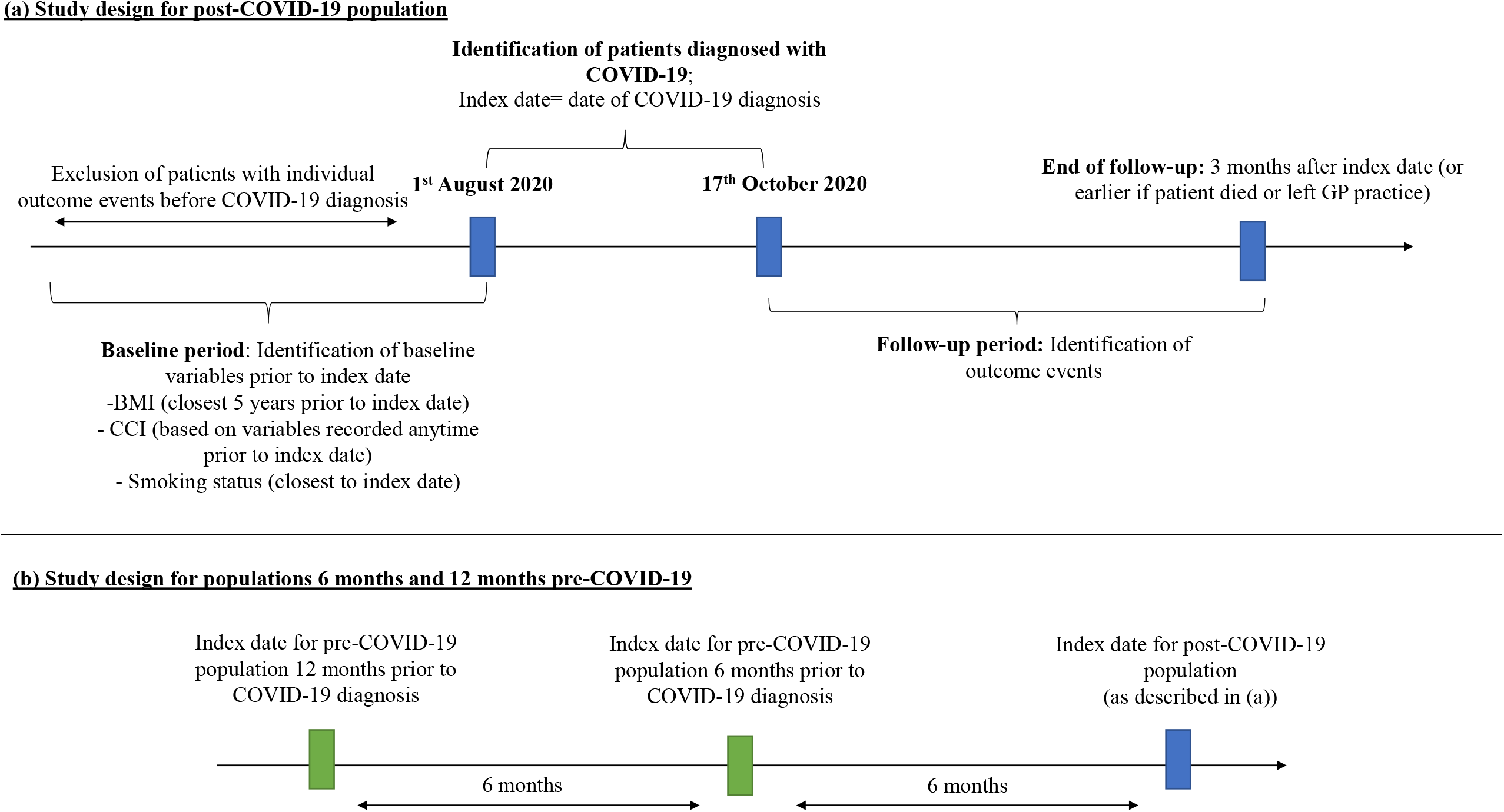
Study design Legend: Follow-up periods for populations 6 months and 12 months pre-COVID-19 were defined the same way as follow-up illustrated in panel (a). For all three populations, maximum follow-up was 3 months post index date.

Patients with evidence of any investigated outcome preceding their COVID-19 diagnosis were excluded from individual analyses to ensure we captured outcomes due to COVID-19 infection rather than pre-existing factors. Outcomes were also determined for the same cohort at 6 and 12 months prior to each patient’s index date. This enabled evaluation of outcome patterns pre-COVID-19 (12 months prior), at the beginning of the pandemic (6 months prior), and post-COVID-19, in the same patients. This was important as changes in healthcare utilisation during the pandemic have been recognised and may influence outcome events.^19^

Baseline characteristics included the most recent measurement of body mass index (BMI) within five years of index date. Smoking status and the Charlson Comorbidity Index (CCI) were identified at any time point before COVID-19 diagnosis. (Fig.1). CCI was identified using a previously published algorithm.^20^

## Definition of Outcomes

Outcomes were considered as symptoms and diseases most likely to affect patients post-infection, guided by previous literature. Codelists were reviewed by a clinician and are accessible at: https://github.com/NHLI-Respiratory-Epi/code_lists/blob/main/25_Long_Covid/Long_Covid_codelists.

Specifically, we identified new symptoms, diseases, prescriptions and healthcare utilisation. New symptoms were chosen in concordance with the NICE 2020 guideline on common symptoms persisting post-acute infection.^10^ For symptom, prescription and healthcare utilisation outcomes, a new event was defined as the first occurrence four weeks after COVID-19 diagnosis, in line with the current NICE definition. For disease outcomes, a new event was defined as the first occurrence after COVID-19 diagnosis. Patients with any of the defined symptoms in the preceding month prior to COVID-19 diagnosis were excluded from analyses. For diseases and prescriptions, a 12-month exclusion window was applied. (Fig. 1). The list of outcomes studied can be found in the appendix (p 3).

### Statistical Analysis

Baseline characteristics are presented as frequencies (%) for categorical data and median with interquartile ranges [IQR] for numerical variables. Event rates and 95% confidence intervals for

each outcome were calculated as the number of patients who experienced the outcome divided by person-time at risk. To understand which patients may be at higher risk of post-COVID-19 outcomes in those managed in the community, event rates were stratified by sex and age (above and below 50 years old).

Cox regression analysis was performed to compare outcome event rates between hospitalised and non-hospitalised cohorts. Finally, we examined the frequency of ten outcomes (anxiety, breathlessness, fatigue, muscle pain, chest pain, chest tightness, insomnia, palpitations, lung fibrosis, and bronchodilator prescription) among the non-hospitalised subgroup. The ten outcomes were those for which there was evidence of an increase in event rates post-COVID-19 relative to prior timepoints. All statistical analyses were conducted using Stata 16. Graphs were created using R 4.0.3.

### Sensitivity analyses

We extended the two-week window for identifying hospitalised COVID-19 cases to three and four weeks, respectively. A further analysis identified symptom and prescription outcomes two weeks after COVID-19 diagnosis instead of four weeks.

### Patient and public involvement

While there has not been any specific patient or user group involvement in the design of this study, patient involvement will be facilitated in communication of findings to patients in long-COVID clinics to guide their ongoing management and care.

### Ethics Approval

A protocol for this research was approved by the Independent Scientific Advisory Committee (ISAC) for MHRA Database Research (protocol number 121_000370) and the approved protocol was made available to the reviewers during peer review. Generic ethical approval for observational research using the CPRD with approval from ISAC has been granted by a Health Research Authority (HRA) Research Ethics Committee (East Midlands – Derby, REC reference number 05/MRE04/87).

## Results

Between August and October 2020, 46,687 patients were diagnosed with COVID-19, of which 45,272 were not hospitalised and 1,415 required hospitalisation. Baseline characteristics are summarised in Table 1. Those hospitalised were older (median age [IQR]: 60 [46 to 74] v. 37 [24 to 53] years), were more likely to be men (52.2% v. 45.2%), overweight or obese (63.1% v 38.9%) and ex- or current smokers (70.8% v. 51.7%) than those not hospitalised. Although hospitalised patients were more likely to have comorbidities, the median number of comorbidities was low in both groups. The geographic distribution of patients in the cohort spanned across England, with the largest patient numbers from the North West (41.5%), London and the South East (14.2%) and the West Midlands (13.9%).(Appendix Table S1).

**Table 1.** Baseline cohort characteristics

|  | **Community**<br>**COVID-19**<br>**N=45,272** | **Hospitalised with**<br>**COVID-19**<br>**N=1,415** | **Total**<br>**N=46,687** |
| --- | --- | --- | --- |

|  | **n (%)** | **n (%)** | **N (%)** |
| --- | --- | --- | --- |
| Male | 20,460 (45.2) | 738 (52.2) | 21,198 (45.4) |
| Female | 24,812 (54.8) | 677 (47.8) | 25,489 (54.6) |
| **Age** |  |  |  |
| 18-30 | 17,928 (39.6) | 116 (8.2) | 18,044 (38.6) |
| 31-40 | 7,602 (16.8) | 150 (10.6) | 7,752 (16.6) |
| 41-50 | 7,127 (15.7) | 187 (13.2) | 7,314 (15.7) |
| 51-60 | 7,206 (15.9) | 260 (18.4) | 7,466 (16) |
| 61-70 | 3,204 (7.1) | 243 (17.2) | 3,447 (7.4) |
| 71-80 | 1,281 (2.8) | 156 (18.1) | 1,537 (3.3) |
| >80 | 924 (2) | 203 (14.3) | 1,127 (2.4) |
| **BMI (kg/m<sup>2</sup>)** |  |  |  |
| ≤18.5 | 889 (2) | 26 (1.8) | 915 (2) |
| 18.5-24.5 | 11,071 (24.5) | 246 (17.4) | 11,317 (24.2) |
| 25.0-29.9 | 9,120 (20.1) | 353 (24.9) | 9,473 (20.3) |
| ≥ 30.0 | 8,526 (18.8) | 541 (38.2) | 9,067 (19.4) |
| Unknown | 15,666 (34.6) | 249 (17.6) | 15,915 (34.1) |
| **Smoking status** |  |  |  |
| Current smoker | 16,019 (35.4) | 619 (43.7) | 16,638 (35.6) |
| Ex-smoker | 7,374 (16.3) | 384 (27.1) | 7,758 (16.6) |
| Never smoked | 19,058 (42.1) | 396 (28) | 19,454 (41.7) |
| Unknown | 2,821 (6.2) | 16 (1.1) | 2,837 (6.1) |
| **CCI** |  |  |  |
| 0 | 28,515 (63) | 415 (29.3) | 28,930 (62) |
| 1-2 | 13,287 (29.3) | 494 (34.9) | 13,781 (29.5) |
| 3-4 | 2,499 (5.5) | 299 (21.1) | 2,798 (6) |
| 5-6 | 742 (1.6) | 139 (9.8) | 881 (1.9) |
| >6 | 229 (0.5) | 68 (4.8) | 297 (0.6) |
| Abbreviations |  |  |  |
| BMI= Body mass index; CCI= Charlson Comorbidity index. |  |  |  |

### Post-COVID-19 outcomes in hospitalised and community patients

Median follow-up time was 63 days [IQR 63 to 63] for determining new symptoms and prescriptions post COVID-19 diagnosis and 91 days [IQR 91 to 91] for new diseases.

### Symptoms

For most symptoms, hospitalised patients had higher event rates than the community group (Fig. 2). These included breathlessness, cough, joint pain, chest pain, fatigue, abdominal pain, nausea, skin rashes, dizziness, fever, diarrhoea, cognitive impairment, and delirium. The largest differences between hospitalised and community patients were respectively noted for rates per 100,000 person-weeks [95%CI] of *breathlessness:* 536 [432 to 663] v. 85 [77 to 93]; *joint pain:* 295 [221 to 392] v. 168 [158 to 179]; *cough:* 150 [101 to 224] v. 50 [44 to 56]; *chest pain:* 157 [107 to 231] v. 50 [44 to 56]; and *fatigue:* 102 [63 to 163] v. 44 [39 to 50], with smaller absolute differences for the remaining eight symptoms (Appendix Table S2).

**Fig. 2:**
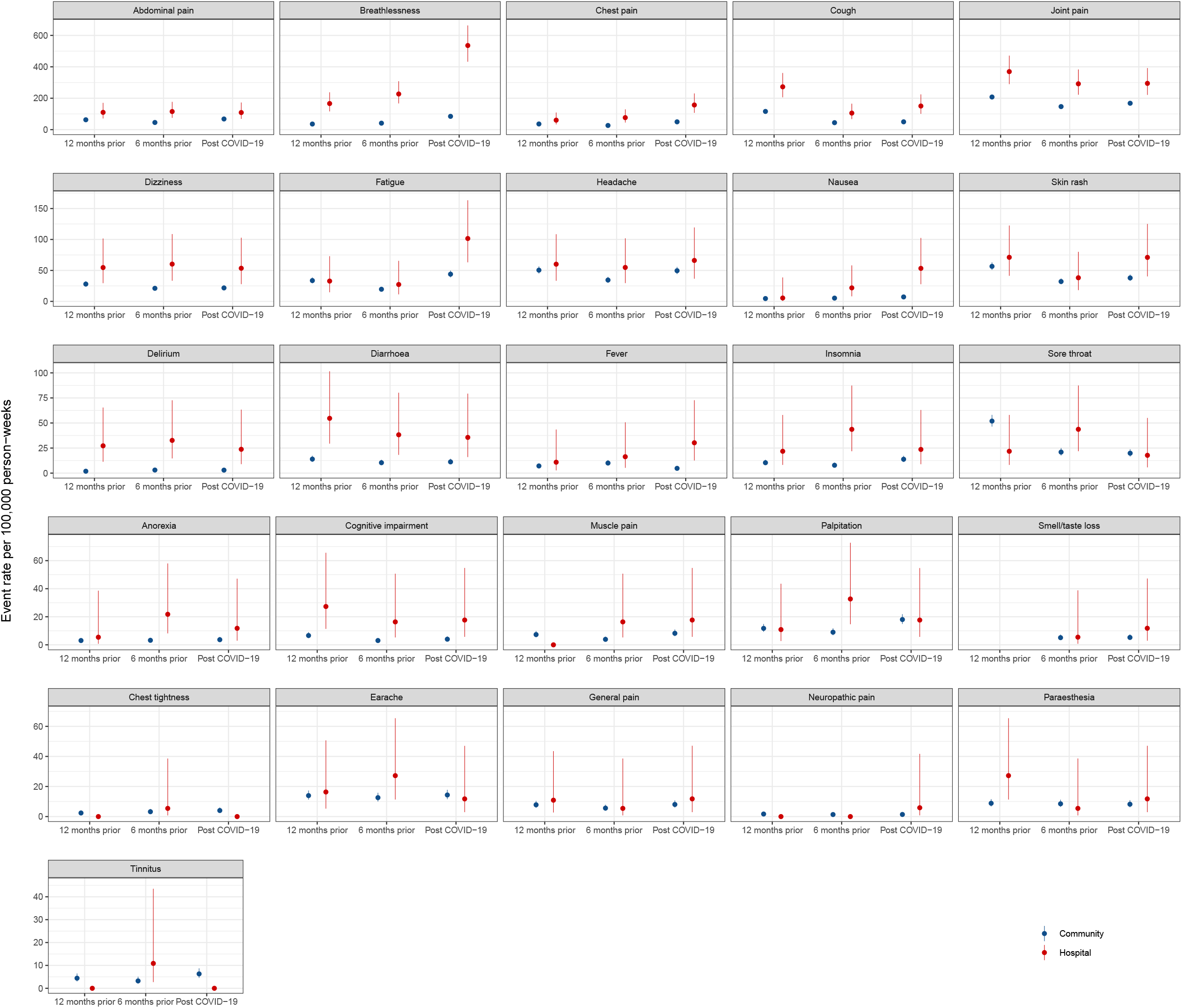
Symptom event rates per 100,000 person-weeks in hospitalised and community COVID-19 patient groups Legend: Vertical bars for each data point show 95% confidence intervals.

There were non-significant differences of general and neuropathic pain, muscle pain, headache, paraesthesia, insomnia, ear/nose/throat symptoms and anorexia (Appendix Table S5).

Palpitation rates were the same in both groups post-COVID-19 but higher rates of chest tightness and tinnitus were noted in the community group, although absolute rates were very low (<10 per 100 000 person weeks) (Appendix Table S2).

### Diseases

Regarding most diseases, hospitalised patients had higher event rates than the community group (Fig. 3). These included all cardiovascular and haematological conditions, diabetes, adrenal disease, renal failure and arthritis. The largest differences between hospitalised and community patients were respectively noted for rates per 100,000 person-weeks [95%CI] of *diabetes*: 303 [225 to 416] v. 36 [32 to 42]; *hypertension:* 244 [178 to 344] v. 47 [41 to 53]; *venous thromboembolism* (*VTE*): 185 [130 to 271] v. 10 [8 to 13]; *renal failure:* 149 [101 to 230] v. 11 [9 to 14] and *anaemia*: 87 [52 to 155] v. 15 [12 to 18] (Appendix Table S3). Rates of asthma, lung fibrosis, GORD, liver disease, anxiety and depression were not statistically significantly different between groups (Appendix Table S3 and S6).

**Fig. 3:**
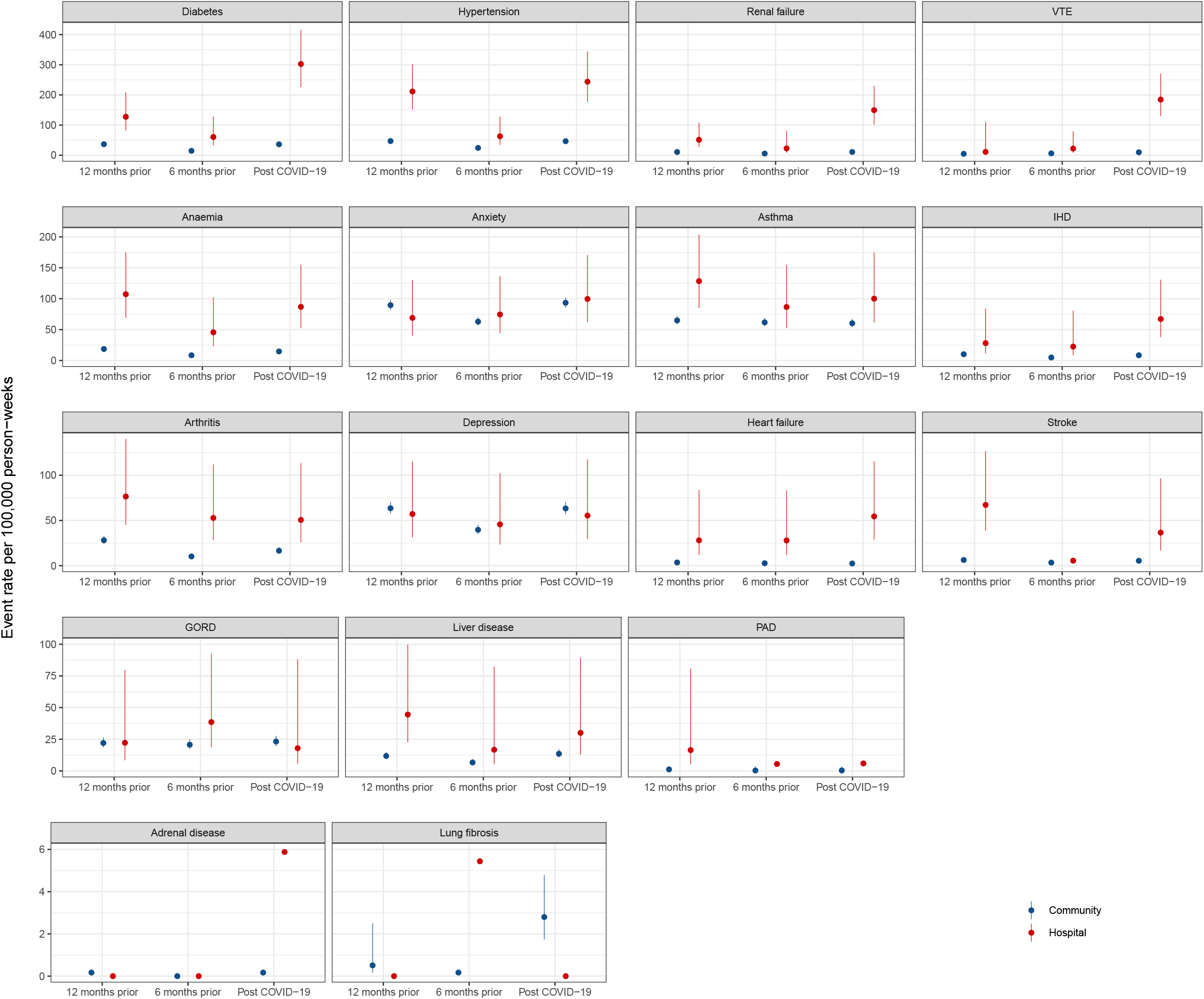
Disease event rates per 100,000 person-weeks in hospitalised and community COVID-19 patient groups Legend: Vertical bars for each data point show 95% confidence intervals. GORD: gastrooesophageal reflux disease; IHD: ischaemic heart disease; PAD: peripheral arterial disease; VTE: venous thromboembolism.

By contrast, absolute rates of lung fibrosis were higher in the community group, although rates were very low (<5 per 100,000 person-weeks). There were no events for either thyroid disease or inflammatory bowel disease in either group.

### Prescriptions

Prescription of all medications occurred more commonly in the hospitalised than the community group post-COVID-19 (Fig. 4). For analgesics, rates respectively per 100,000 person-weeks [95%CI] were for opiates 378 [282 to 508] v. 82 [74 to 90]; paracetamol 304 [226 to 411] v. 19 [16 to 23]; and NSAIDs 303 [221 to 414] v. 84 [76 to 92] (Appendix Table S4 and S6). Inhalers such as bronchodilators (212 [147 to 308]) were prescribed more frequently than inhaled corticosteroids (144 [93 to 224]) in the hospitalised group. By contrast, diuretics were prescribed less frequently than other medications.

**Fig. 4:**
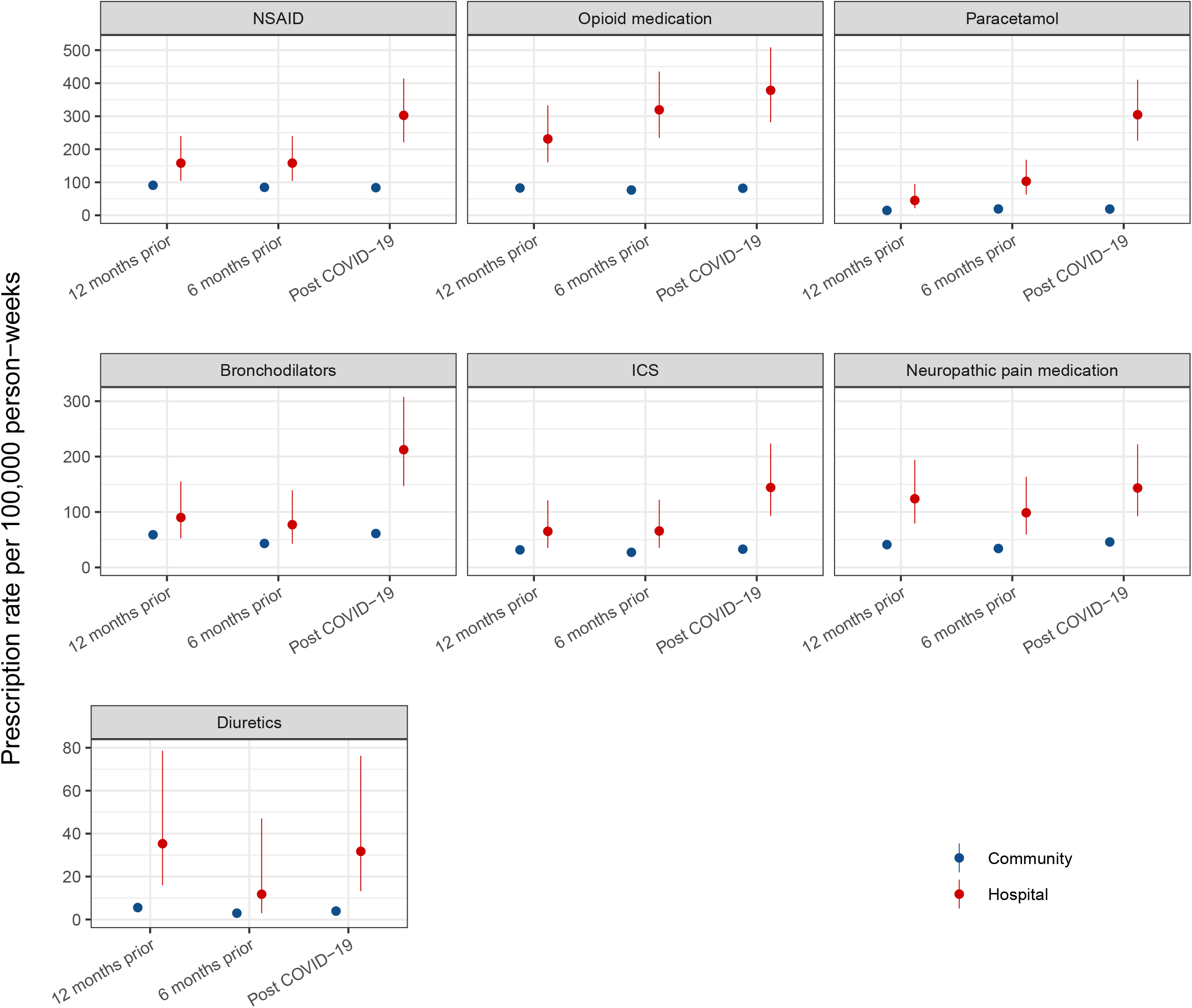
Prescription event rates per 100,000 person-weeks in hospitalised and community COVID-19 patient groups Legend: Vertical bars for each data point show 95% confidence intervals. NSAID: non-steroidal anti-inflammatory drugs; ICS: inhaled corticosteroids.

### Healthcare utilisation

The hospitalised group utilised more healthcare (including GP visits, referrals, A&E, hospitalisation) than the community group post-COVID-19, with a 2.7-fold difference in rates per 100,000 person-weeks [95%CI] between groups (52,775 [50,570 to 55,105] v. 19,405 [19,142 to 19,673]) in hospitalised and community groups, respectively (Appendix Table S5).

### Post-COVID-19 outcomes relative to the 6 and 12 months pre-COVID-19

#### Symptoms

Both groups experienced an increase in rates for chest pain, fatigue and breathlessness post-COVID-19 relative to 12 months prior (Fig. 2, Appendix Table S2). Rates increased for some symptoms for only those hospitalised post-COVID-19 relative to 12 months prior, with nausea showing the largest increase. Although absolute rates were very low, small increases in rates of chest tightness, tinnitus, insomnia and palpitations over the same period were only noted in the community group. For both groups, rates of cough were lower post-COVID-19 relative to pre-pandemic rates, while general and abdominal pain, headache, delirium, dizziness and earache remained similar.

#### Diseases

Lower rates of hypertension, stroke and diabetes were noted at 6 months prior relative to other timepoints in both groups (Fig. 3, Appendix Table S3). While rates of renal failure, diabetes and adrenal disease remained relatively stable in the community group post-COVID-19 relative to 12 months prior, rates of renal failure and diabetes increased 2.92 and 2.37 fold, respectively, in the hospitalised group. VTE rates increased in both groups, but significantly more in the hospitalised group. By contrast, rates of depression were very similar post-COVID-19 relative to those pre-pandemic in both groups.

#### Prescriptions

At all timepoints, those hospitalised had higher prescription rates for each medication type than the community group (Fig. 4). Rates of diuretics were broadly similar post-COVID-19 in both groups relative to pre-pandemic rates (Appendix Table S4). While rates for inhalers and analgesics remained similar over the same time period for the community group, rates increased for those hospitalised (bronchodilators: 2.35 fold; ICS 2.2 fold; paracetamol 6.75 fold; NSAIDs 1.9 fold; opiates 1.64 fold).

#### Healthcare utilisation

Differences in healthcare utilisation post-COVID-19 between the two groups showed a similar pattern to those at both prior timepoints. However, while healthcare utilisation increased in both groups post-COVID-19 relative to pre-pandemic levels, this was much higher in the hospitalised group (61.2% increase v. 28.5%). Healthcare utilisation was lower 6 months prior relative to other timepoints for each group (Appendix Table S5).

#### Stratification of outcomes by age and sex in community group

Overall, men <50 had the lowest rates of symptoms, disease, and prescriptions. There were differences in age whereby older adults had higher rates of breathlessness, chest pain, cognitive impairment, dizziness, delirium, muscle pain, cough, diabetes, arthritis, VTE, and opioid, paracetamol, and diuretic prescriptions compared to younger adults. Older adults also had higher rates of cardiovascular disease, notably in men.

Women had higher rates of fatigue and older women in particular had higher rates of joint pain compared to men. Younger women had higher rates of headache and anxiety compared to men and higher rates of skin rash, depression, and sore throat compared to men and older adults. (Fig. 5-7, Appendix Tables S7-S18).

**Fig. 5:**
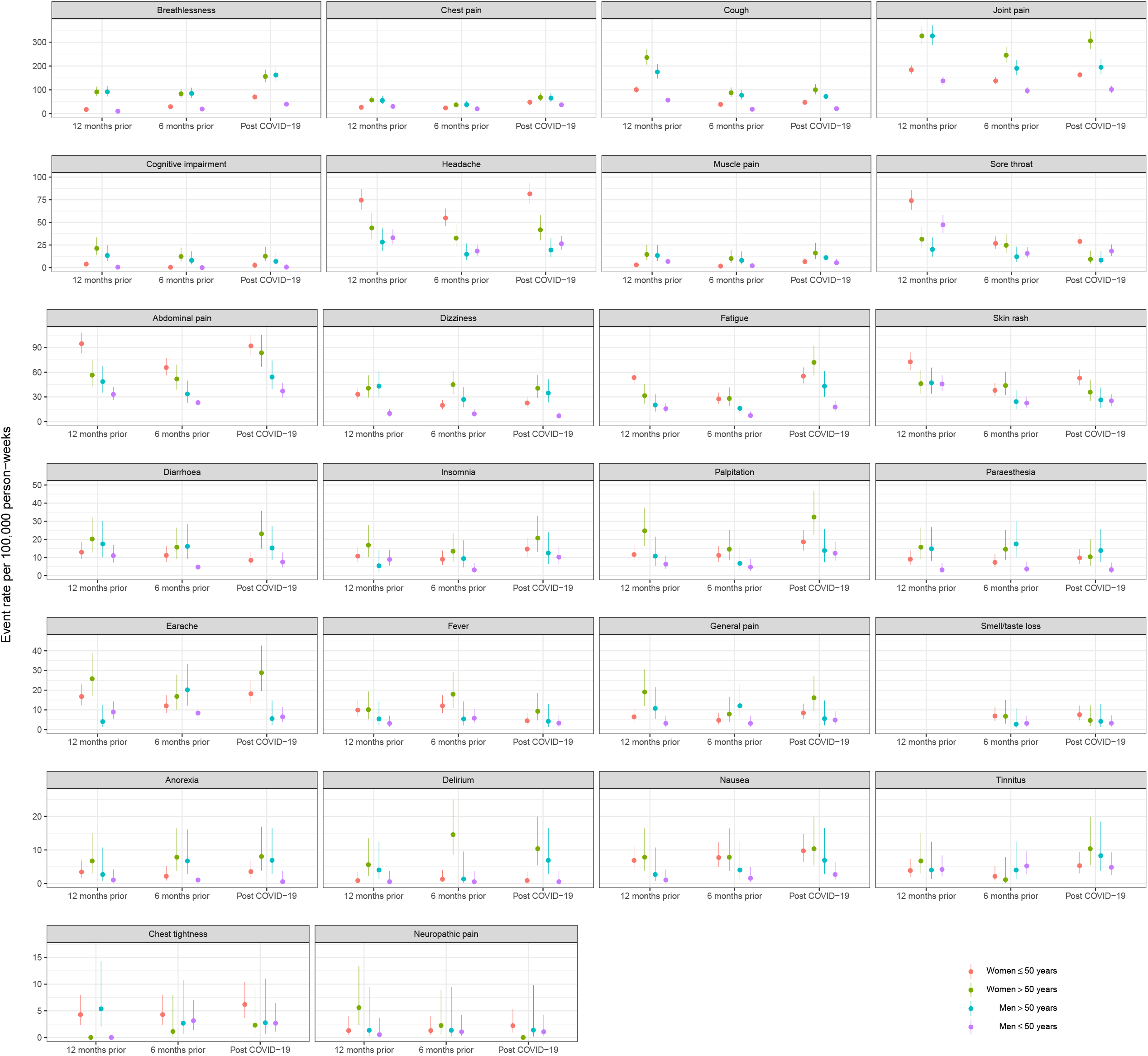
Symptom event rates per 100,000 person-weeks in patients with community COVID-19 stratified by age and sex. Legend: Vertical bars for each data point show 95% confidence intervals.

**Fig. 6:**
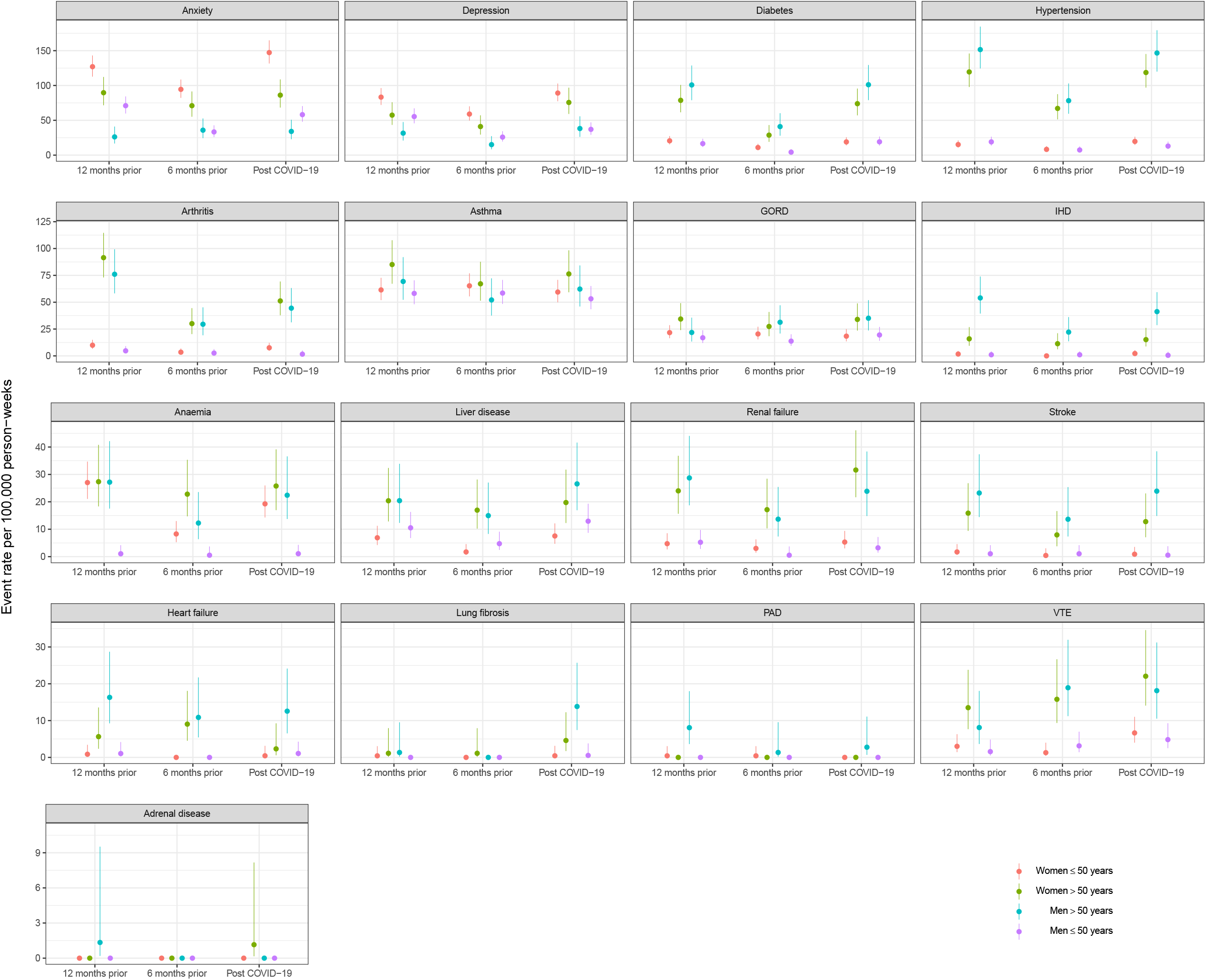
Disease event rates per 100,000 person-weeks in patients with community COVID-19 stratified by age and sex. Legend: Vertical bars for each data point show 95% confidence intervals. GORD: gastrooesophageal reflux disease; IHD: ischaemic heart disease; PAD: peripheral arterial disease; VTE: venous thromboembolism.

**Fig. 7:**
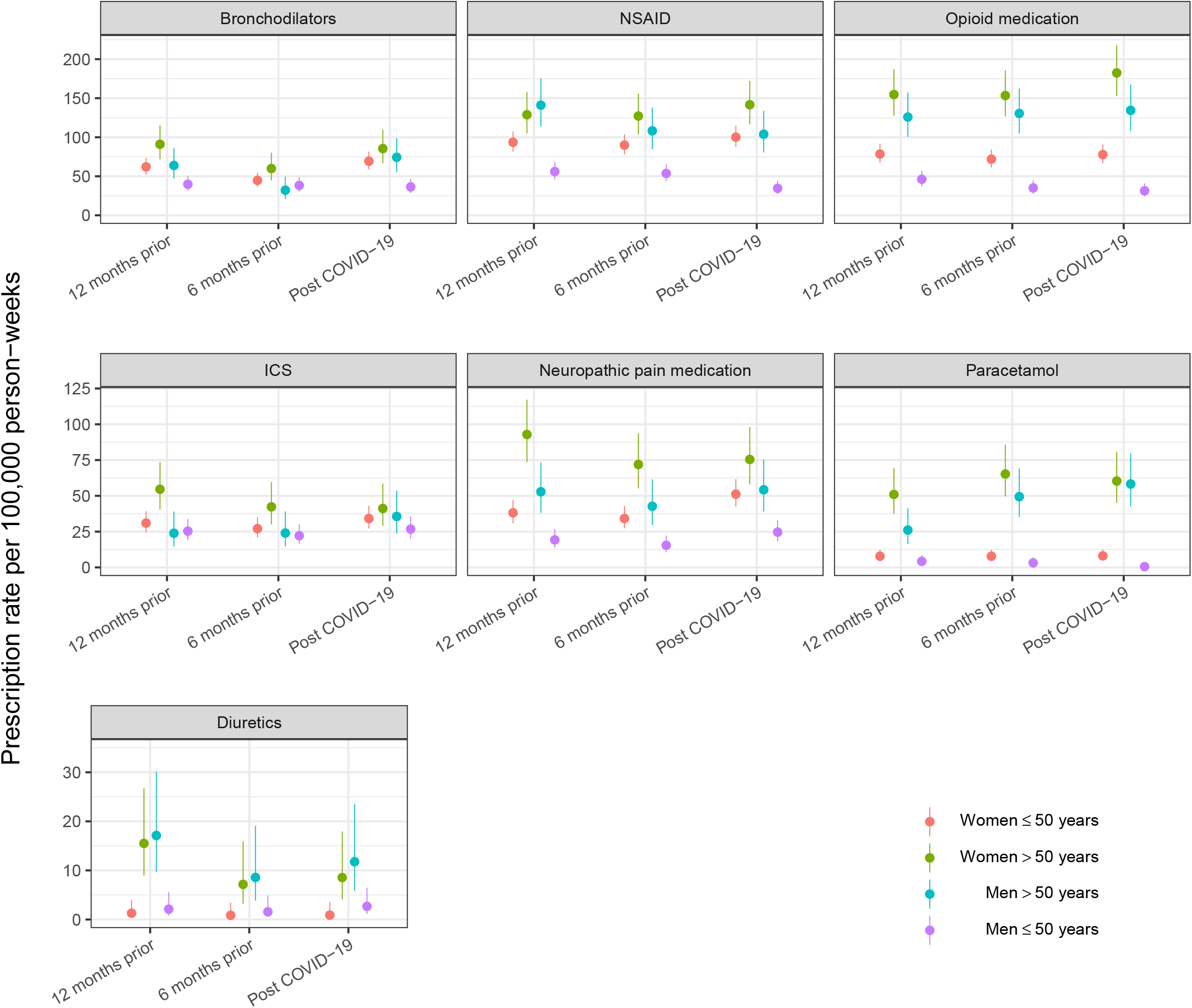
Prescription event rates per 100,000 person-weeks in patients with community COVID-19 stratified by age and sex. Legend: Vertical bars for each data point show 95% confidence intervals. NSAID: non-steroidal anti-inflammatory drugs; ICS: inhaled corticosteroids.

#### Co-occurrence of outcomes post-COVID-19 in community group

The majority of the community COVID-19 population (43,390 of 45,272 [95.8%]) did not experience any of the ten outcomes identified as having increased event rates post infection relative to the 6 and 12-month prior periods. A total of 1,882 (4.2%) individuals experienced at least one of these outcomes. Anxiety, breathlessness, chest pain, fatigue, and bronchodilator prescriptions were the most frequent outcomes. Approximately 25% of patients experienced anxiety or breathlessness, 15% chest pain, 13.3% fatigue, and 16.7% received a new bronchodilator prescription, of which 90.3% were short acting beta-agonists.

#### Sensitivity analysis

Classification of patients as hospitalised cases at three or four weeks yielded similar results to using a two week window. The number of patients classed as hospitalised cases within three weeks of a positive test was 1,633 (3.5%); within four weeks this increased to 1,814 (3.9%). Analyses of event rates that identified symptom and prescription outcomes from two weeks after COVID-19 diagnosis showed similar results to those described in figures 2-4. (appendix Tables S19-S22)

## Discussion

### Principal Findings

This is the first population-based study in the UK investigating symptoms, diseases, prescriptions and healthcare utilisation among people hospitalised or managed in the community following COVID-19 diagnosis. For most outcomes, hospitalised patients had higher event rates than the community group. However, there were pertinent differences between groups, with the post-COVID-19 community group having higher rates of chest tightness, tinnitus and lung fibrosis, thus highlighting important differences in post-COVID-19 sequelae and suggesting that post-COVID-19 follow up and management strategies will need to be tailored appropriately.

Further in-keeping with the lower rate of outcomes in those managed in the community in comparison to those hospitalised, a post-acute burden of sequelae was captured in only 4.2% of cases managed in the community. Anxiety, breathlessness, chest pain and fatigue were most frequently reported, alongside prescriptions for bronchodilators. Furthermore, age and sex-specific differences in outcomes were noted, with older adults experiencing higher rates of most symptoms, cardiovascular disease, diabetes, VTE and analgesic prescriptions such as paracetamol and opiates. In addition, women had higher rates of fatigue, with higher rates of headache, anxiety and depression in younger women. While primary care consultation rates have been shown to be lower in men than women and may account for some of the sex-specific differences noted, consultation rates in men and women with comparable underlying morbidities tend to be similar.^21^

While healthcare utilisation increased in both groups post-COVID-19 relative to pre-pandemic levels, this increase was significantly higher in the hospitalised group. Nevertheless, healthcare utilisation in the community group increased 28.5% in the post-COVID-19 period relative to pre-pandemic levels, highlighting a need to ensure adequate provision of care for this population. This increase since pre-pandemic levels also suggests that lack of access to healthcare following the second wave is less of a problem than in wave one; however, evidence indicates this can nevertheless still be difficult.^16,19^ Contrary to our findings however, a Norwegian study suggested that mild COVID-19 does not persist to cause a need for healthcare beyond two months following a positive test.^22^

It is unclear if symptoms or diseases post-COVID-19 are due to the infection itself, anxiety caused by diagnosis and isolation or due to complications. Whilst this study does not inform us about mechanisms, it further paints the picture, helping us understand the population-level burden of post-COVID-19 sequelae across the spectrum of acute infection severity.

### Comparison with related studies

There is a growing body of literature on post-acute COVID-19 outcomes among people who have been hospitalised, ^23-24^ but little that compares those managed in hospitals with those managed in the community. Our findings are in keeping with the few such studies that exist, whilst adding a depth of understanding of the situation in the UK. A study from Israel found a substantially lower prevalence of ongoing issues in the community population compared with hospitalised cohorts in other studies, the most common being fatigue, myalgia, runny nose and shortness of breath.^25^ Small studies investigating lung function in the non-hospitalised post-acute COVID-19 population have also found that over two months post infection, 46% of patients remained symptomatic, with lower lung function in this group than asymptomatic patients.^26^ A US-based study comparing post-acute COVID-19 sequelae in those with mild, moderate and severe illness found older age groups were more likely to report persistent symptoms, the most common being fatigue (13·6%) and anosmia (13·6%).^27^ Another study from the Netherlands and Belgium found a larger median reduction in symptom number at 80 days follow-up in those non-hospitalised compared with those hospitalised.^15^ However, only 0.7% of their respondents were symptom-free at 80 days, highlighting that surveys and healthcare record data do provide different magnitudes of effect. Indeed, the aforementioned Israeli study which used both methods found differences in prevalence between self-reported symptoms and health records.^25^ Whilst survey estimates of symptom burden in those non-hospitalised are much higher than in our study, this is most likely due to selection bias, coupled with a higher tendency towards symptom reporting in surveys and in those with suspected COVID-19.^15, 25^ Persistent fatigue in both those hospitalised and not has been a common finding in several studies.^3, 25, 27^

In common with other studies, we found high rates of post-COVID-19 diabetes in the hospitalised^24^; diabetes was also one of the diseases with the largest hospital-community differences. A bidirectional relationship has been postulated between diabetes and COVID-19, with diabetes leading to higher risk of infection and worse outcomes but also some evidence that COVID-19 may precipitate diabetes.^28^ This is complicated by the fact there has been a reported reduction in diagnosis of type 2 diabetes during the pandemic.^29^ Likewise, renal impairment and VTE have been recognised post hospitalisation.^7,30^

### Strengths and limitations

Our study purposefully only included COVID-19 patients from wave two, when testing capacity was much higher, thereby limiting potential selection biases. That our proportion of hospitalised patients is in keeping with UK national estimates increases our confidence that our study population is broadly representative. Whilst we cannot ascertain whether symptoms recorded on primary care records were directly due to COVID-19 or other conditions, we did investigate event rates among the same cohorts 6 and 12 months prior to contextualize our findings. In addition, we did include a window of no symptoms to ensure diseases, symptoms and prescriptions captured were new and not pre-existing to the COVID-19 diagnosis.

Risk of misclassification of diseases and symptoms, an inherent weakness of studies of this nature, was minimised by using previously validated codelists wherever possible and creating codelists tailored to the objectives of this study. We were unable to capture the effect of socioeconomic status as these data are not available in primary care records and accept that this is a limitation. Nor have we been able to consider the severity of our investigated outcomes.

Given the relatively short follow-up period, we may be missing some symptoms and diseases which occur later in the trajectory of long-Covid. For this reason, we plan to repeat this analysis in the future. It is also likely that we will have missed some cases of new onset symptoms or diseases post-COVID in those patients who choose not to seek medical care and manage symptoms independently with over-the-counter medications. Furthermore, qualitative studies have revealed perceived barriers in primary care accessibility and limited understanding of long-Covid among clinicians.^16^ These factors may also lead to underreporting and under-recording of symptoms/diseases.

### Implications of our findings

With over 4 million individuals recovered from acute COVID-19 in the UK at the time of writing,^13^ our findings of multi-system symptom and disease burden, with a predominance in those hospitalised, has significant implications for future healthcare service planning. While healthcare utilisation increased relatively more in those hospitalised, there was nevertheless an increase in those managed in the community too, suggesting current healthcare service provision may need to be moulded to meet this increasing demand. Furthermore, given the significant interest in long-Covid globally and calls for further research in assessing interventions in individuals continuing to suffer ongoing effects of COVID-19 who have not been hospitalised, findings from this study are timely. To date, the focus has overwhelmingly been on studies assessing outcomes in hospitalised COVID-19 patients, with a significant paucity of studies exploring outcomes in those with milder disease who did not require hospitalisation. The multi-system nature of symptoms and diseases noted in both non-hospitalised and hospitalised cases highlights the importance of providing integrated multidisciplinary care in post-COVID-19 management.

## Conclusions

We have shown that across symptom, disease, prescription and healthcare utilisation outcomes, people who were hospitalised with COVID-19 had higher event rates in the post-acute phase compared with those managed in the community. Among the latter, post-acute COVID-19 outcomes were higher in those over 50 years old and in women. Of those managed in the community with any post-acute burden, anxiety, breathlessness, chest pain and fatigue were the most frequently reported. Studies with longer follow-up will help provide further insight into the longitudinal trajectory of post-COVID-19 sequelae, however it appears that ongoing needs in those admitted to hospital or managed in the community will differ.

#### What is already known on this topic

- Persistence of symptoms and new organ dysfunction post-COVID 19 have been recognised in several small observational studies but this has been primarily in hospitalised patients with more severe disease.
- Few studies compare outcomes between non-hospitalised and hospitalised individuals, with studies to date limited by small cohort sizes and selection biases; no large population-based cohort studies exist assessing outcomes between groups.

#### What this study adds

- Hospitalised patients had higher rates of most post-COVID-19 sequelae compared to those managed in the community, with additional age and sex-specific differences in outcome rates for the latter.
- Healthcare utilisation in the community group increased 28.5% post-COVID-19 relative to pre-pandemic. However, only a small proportion (4.2%) of this group experienced outcomes resulting in increased event rates post infection relative to pre-diagnosis; anxiety, breathlessness, chest pain and fatigue were most frequently reported.
- This large population-based study complements ongoing work exploring outcomes post-COVID-19, while also highlighting that post-COVID-19 management strategies will need to be tailored to specific patient group needs.

## Data Availability

Linked pseudonymised mortality data from the Office for National Statistics (ONS), socioeconomic data from the Index of Multiple Deprivation (IMD), and secondary care data from Hospital Episode Statistics (HES) were provided for this study by CPRD for patients in England. Data is linked by NHS Digital, the statutory trusted third party for linking data, using identifiable data held only by NHS Digital. Select general practices consent to this process at a practice level, with individual patients having the right to opt-out. Use of HES and ONS data is Copyright (2018), re-used with the permission of The Health & Social Care Information Centre, all rights reserved.
Data are available on request from the CPRD. Their provision requires the purchase of a license, and this license does not permit the authors to make them publicly available to all. This work used data from the version collected in January 2021 and have clearly specified the data selected in the Methods section. To allow identical data to be obtained by others, via the purchase of a license, the code lists have been provided on GitHub. Licenses are available from the CPRD (http://www.cprd.com): The Clinical Practice Research Datalink Group, The Medicines and Healthcare products Regulatory Agency, 10 South Colonnade, Canary Wharf, London E14 4PU.

## Contributors

JKQ conceptualised the study and all authors contributed to study design. HRW, CG, AK, AM, CI, and RS created codelists for outcomes of interest. HRW, CG, CK and AM prepared the data and performed statistical analyses, with all authors contributing to interpretation of results. Figures were created by CG. AK, HRW, AM, CG and JKQ drafted the original manuscript, with critical revision of the manuscript by all authors. All authors approved the final manuscript. The corresponding author attests that all listed authors meet authorship criteria and that no others meeting the criteria have been omitted. The corresponding author is also the guarantor for this manuscript and accepts full responsibility for the work, had access to all the data and was responsible for the decision to publish.

## Funding

This work is supported by BREATHE - The Health Data Research Hub for Respiratory Health [MC_PC_19004]. BREATHE is funded through the UK Research and Innovation Industrial Strategy Challenge Fund and delivered through Health Data Research UK. The funder had no role in study design, data collection, analysis or interpretation, or manuscript writing. All authors had full access to all the data in the study and had final responsibility for the decision to submit for publication.

## Declaration of interests

HRW, CG, AK, CK, AM, CI, MW have nothing to declare. RG is a current employee of Gilead Sciences, outside the submitted work. JKQ reports grants from AUK-BLF, The Health Foundation, grants and personal fees from AZ, BI, GSK, Bayer, grants from Chiesi, outside the submitted work.

## Acknowledgements

This research was supported by the National Institute for Health Research (NIHR) Imperial Biomedical Research Centre (BRC). The views expressed are those of the authors and not necessarily those of the NIHR or the Department of Health and Social Care.

## Ethics and Disclosure Statement

This work is based on data from the Clinical Practice Research Datalink (CPRD) obtained under license from the United Kingdom (UK) Medicines and Healthcare products Regulatory Agency (MHRA). The data is provided by patients and collected by the National Health Service (NHS) as part of their care and support. The interpretation and conclusions contained in this study are those of the authors alone.

## Transparency statement

The corresponding author confirms that the manuscript is an honest, accurate, and transparent account of the study being reported, that no important aspects of the study have been omitted and that any discrepancies from the study as originally planned have been explained.

## Data Availability

Linked pseudonymised mortality data from the Office for National Statistics (ONS), socioeconomic data from the Index of Multiple Deprivation (IMD), and secondary care data from Hospital Episode Statistics (HES) were provided for this study by CPRD for patients in England. Data is linked by NHS Digital, the statutory trusted third party for linking data, using identifiable data held only by NHS Digital. Select general practices consent to this process at a practice level, with individual patients having the right to opt-out. Use of HES and ONS data is Copyright © (2018), re-used with the permission of The Health & Social Care Information Centre, all rights reserved.

Data are available on request from the CPRD. Their provision requires the purchase of a license, and this license does not permit the authors to make them publicly available to all. This work used data from the version collected in January 2021 and have clearly specified the data selected in the Methods section. To allow identical data to be obtained by others, via the purchase of a license, the code lists have been provided on GitHub. Licenses are available from the CPRD (http://www.cprd.com): The Clinical Practice Research Datalink Group, The Medicines and Healthcare products Regulatory Agency, 10 South Colonnade, Canary Wharf, London E14 4PU.

## Data sharing

This study used existing data from the UK CPRD electronic health record database, this data resource is accessible only to researchers with protocols approved by the CPRD’s independent scientific advisory committee; therefore, no additional unpublished data are available. All data management and analysis computer code are available on request. The study protocol and analysis plan are available in the associated supplementary material.

## License statement for publication

The Corresponding Author has the right to grant on behalf of all authors and does grant on behalf of all authors, a worldwide license to the Publishers and its licensees in perpetuity, in all forms, formats and media (whether known now or created in the future), to i) publish, reproduce, distribute, display and store the Contribution, ii) translate the Contribution into other languages, create adaptations, reprints, include within collections and create summaries, extracts and/or, abstracts of the Contribution, iii) create any other derivative work(s) based on the Contribution, to exploit all subsidiary rights in the Contribution, v) the inclusion of electronic links from the Contribution to third party material where-ever it may be located; and, vi) license any third party to do any or all of the above.

## Notes

### Competing Interest Statement

The authors have declared no competing interest.

### Clinical Trial

ISAC approval obtained from CPRD

### Author Declarations

A protocol for this research was approved by the Independent Scientific Advisory Committee (ISAC) for MHRA Database Research (protocol number 121_000370) and the approved protocol was made available to the reviewers during peer review. Generic ethical approval for observational research using the CPRD with approval from ISAC has been granted by a Health Research Authority (HRA) Research Ethics Committee (East Midlands Derby, REC reference number 05/MRE04/87).

