## Supplementary tables and figures for "Post-acute COVID-19 sequelae in cases managed in the community or hospital in the UK: a population based study"

**Supplementary material**

**List of abbreviations**

**IHD** - Ischemic Heart Disease
**PAD** - Peripheral Arterial Disease
**VTE** - Venous Thromboembolism
**GORD** - Gastro-Oesophageal Reflux Disease
**ICS** - Inhaled Corticosteroids
**NSAIDS** - Non-Steroidal Anti-Inflammatory Drugs

### List of new symptoms, non-communicable diseases, medication prescriptions, and definition of healthcare utilisation

1. **New symptoms** included: respiratory (breathlessness and cough), cardiovascular (chest tightness, chest pain, palpitations), gastrointestinal (abdominal pain, anorexia/reduced appetite, nausea, diarrhoea), musculoskeletal (joint and muscle pain), dermatological (skin rashes), neurological (headache, dizziness, insomnia, cognitive impairment, delirium, paresthesia), ear nose and throat (tinnitus, earache, sore throat, loss of smell) and general symptoms (fatigue, fever, pain).

2. **New non-communicable diseases** including: cardiovascular (hypertension, ischaemic heart disease (IHD), heart failure, peripheral arterial disease (PAD), stroke/transient ischaemic attack) , respiratory (asthma, pulmonary fibrosis), gastrointestinal (inflammatory bowel disease (IBD), liver disease, gastro-oesophageal reflux (GORD)), endocrine (diabetes, thyroid and adrenal disorders), renal failure, rheumatological (arthritis), haematological (venous thromboembolism (VTE), anaemia) and psychiatric disorders (delirium, anxiety, depression).

3. **New medication prescriptions** including bronchodilators, inhaled corticosteroids (ICS), analgesics (paracetamol, non-steroidal anti-inflammatory drugs (NSAIDs), opiates, neuropathic pain agents) and diuretics.

4. **New healthcare utilisation** including number of GP visits and hospital admissions for any cause.

### Table S1: Frequency of patients by geographic location in England

| **Geographic location in England** | **Frequency of patients n(%) N=46,687** |
| --- | --- |
| **North east** | 3,122 (6.7) |
| **North west** | 19,359 (41.5) |
| **Yorkshire and the Humber** | 1,875 (4.0) |
| **East Midlands** | 3,020 (6.5) |
| **West Midlands** | 6,495 (13.9) |
| **East** | 1,025 (2.2) |
| **South west** | 2,691 (5.8) |
| **South central** | 2,356 (5.1) |
| **London** | 5,178 (11.1) |
| **South east coast** | 1,451 (3.1) |
| **Unknown** | 115 (0.3) |

### Table S2: Symptom event rates (per 100,000 person-weeks)

| **Outcome** | **Time point** | **COVID exposure group** | **Number of events** | **Time (person-weeks (PW))** | **Rate (per 100,000 PW)** | **Lower CI** | **Upper CI** |
| --- | --- | --- | --- | --- | --- | --- | --- |
| **General pain** | Post-COVID | Community | 46 | 5.7 | 8 | 6 | 11 |
|  |  | Hospital | <10 | 0.2 | 12 | 3 | 47 |
|  | 6 months prior | Community | 33 | 5.9 | 6 | 4 | 8 |
|  |  | Hospital | <10 | 0.2 | 5 | 1 | 39 |
|  | 12 months prior | Community | 46 | 5.9 | 8 | 6 | 10 |
|  |  | Hospital | <10 | 0.2 | 11 | 3 | 44 |
| **Chest pain** | Post-COVID | Community | 283 | 5.7 | 50 | 44 | 56 |
|  |  | Hospital | 26 | 0.2 | 157 | 107 | 231 |
|  | 6 months prior | Community | 156 | 5.9 | 27 | 23 | 31 |
|  |  | Hospital | 14 | 0.2 | 77 | 45 | 129 |
|  | 12 months prior | Community | 212 | 5.9 | 36 | 32 | 41 |
|  |  | Hospital | 11 | 0.2 | 60 | 33 | 109 |
| **Abdominal pain** | Post-COVID | Community | 386 | 5.7 | 68 | 61 | 75 |
|  |  | Hospital | 18 | 0.2 | 109 | 69 | 173 |
|  | 6 months prior | Community | 267 | 5.9 | 46 | 40 | 51 |
|  |  | Hospital | 21 | 0.2 | 115 | 75 | 177 |
|  | 12 months prior | Community | 367 | 5.8 | 63 | 57 | 70 |
|  |  | Hospital | 20 | 0.2 | 110 | 71 | 171 |
| **Headache** | Post-COVID | Community | 281 | 5.7 | 50 | 44 | 56 |
|  |  | Hospital | 11 | 0.2 | 66 | 37 | 119 |
|  | 6 months prior | Community | 202 | 5.9 | 34 | 30 | 40 |
|  |  | Hospital | 10 | 0.2 | 55 | 29 | 102 |
|  | 12 months prior | Community | 295 | 5.8 | 50 | 45 | 57 |
|  |  | Hospital | 11 | 0.2 | 60 | 33 | 108 |
| **Joint pain** | Post-COVID | Community | 939 | 5.6 | 168 | 158 | 179 |
|  |  | Hospital | 47 | 0.2 | 295 | 221 | 392 |
|  | 6 months prior | Community | 850 | 5.8 | 147 | 137 | 157 |
|  |  | Hospital | 52 | 0.2 | 292 | 222 | 383 |
|  | 12 months prior | Community | 1191 | 5.7 | 208 | 196 | 220 |
|  |  | Hospital | 65 | 0.2 | 369 | 290 | 471 |
| **Muscle pain** | Post-COVID | Community | 47 | 5.7 | 8 | 6 | 11 |
|  |  | Hospital | <10 | 0.2 | 18 | 6 | 55 |
|  | 6 months prior | Community | 23 | 5.9 | 4 | 3 | 6 |
|  |  | Hospital | <10 | 0.2 | 16 | 5 | 51 |
|  | 12 months prior | Community | 43 | 5.9 | 7 | 5 | 10 |
|  |  | Hospital | <10 | 0.2 | 0 | n/a | n/a |
| **Neuropathic pain** | Post-COVID | Community | <10 | 5.7 | 1 | 1 | 3 |
|  |  | Hospital | <10 | 0.2 | 6 | 1 | 42 |
|  | 6 months prior | Community | <10 | 5.9 | 1 | 1 | 3 |
|  |  | Hospital | <10 | 0.2 | 0 | n/a | n/a |
|  | 12 months prior | Community | 10 | 5.9 | 2 | 1 | 3 |
|  |  | Hospital | <10 | 0.2 | 0 | n/a | n/a |
| **Fatigue** | Post-COVID | Community | 250 | 5.7 | 44 | 39 | 50 |
|  |  | Hospital | 17 | 0.2 | 102 | 63 | 163 |
|  | 6 months prior | Community | 115 | 5.9 | 20 | 16 | 24 |
|  |  | Hospital | <10 | 0.2 | 27 | 11 | 65 |
|  | 12 months prior | Community | 197 | 5.9 | 34 | 29 | 39 |
|  |  | Hospital | <10 | 0.2 | 33 | 15 | 73 |
| **Fever** | Post-COVID | Community | 27 | 5.7 | 5 | 3 | 7 |
|  |  | Hospital | <10 | 0.2 | 30 | 13 | 73 |
|  | 6 months prior | Community | 59 | 5.9 | 10 | 8 | 13 |
|  |  | Hospital | <5 | 0.2 | 16 | 5 | 51 |
|  | 12 months prior | Community | 42 | 5.9 | 7 | 5 | 10 |
|  |  | Hospital | <5 | 0.2 | 11 | 3 | 43 |
| **Breathlessness** | Post-COVID | Community | 479 | 5.7 | 85 | 77 | 93 |
|  |  | Hospital | 84 | 0.2 | 536 | 432 | 663 |
|  | 6 months prior | Community | 242 | 5.9 | 41 | 36 | 47 |
|  |  | Hospital | 41 | 0.2 | 227 | 167 | 308 |
|  | 12 months prior | Community | 210 | 5.9 | 36 | 31 | 41 |
|  |  | Hospital | 30 | 0.2 | 166 | 116 | 238 |
| **Cough** | Post-COVID | Community | 281 | 5.6 | 50 | 44 | 56 |
|  |  | Hospital | 24 | 0.2 | 150 | 101 | 224 |
|  | 6 months prior | Community | 259 | 5.8 | 44 | 39 | 50 |
|  |  | Hospital | 19 | 0.2 | 105 | 67 | 165 |
|  | 12 months prior | Community | 676 | 5.8 | 116 | 108 | 125 |
|  |  | Hospital | 49 | 0.2 | 273 | 206 | 361 |
| **Chest tightness** | Post-COVID | Community | 23 | 5.7 | 4 | 3 | 6 |
|  |  | Hospital | <10 | 0.2 | 0 | n/a | n/a |
|  | 6 months prior | Community | 19 | 5.9 | 3 | 2 | 5 |
|  |  | Hospital | <10 | 0.2 | 5 | 1 | 39 |
|  | 12 months prior | Community | 14 | 5.9 | 2 | 1 | 4 |
|  |  | Hospital | <10 | 0.2 | 0 | n/a | n/a |
| **Palpitations** | Post-COVID | Community | 103 | 5.7 | 18 | 15 | 22 |
|  |  | Hospital | <10 | 0.2 | 18 | 6 | 55 |
|  | 6 months prior | Community | 53 | 5.9 | 9 | 7 | 12 |
|  |  | Hospital | <10 | 0.2 | 33 | 15 | 73 |
|  | 12 months prior | Community | 69 | 5.9 | 12 | 9 | 15 |
|  |  | Hospital | <10 | 0.2 | 11 | 3 | 44 |
| **Diarrhea** | Post-COVID | Community | 64 | 5.7 | 11 | 9 | 14 |
|  |  | Hospital | <10 | 0.2 | 36 | 16 | 79 |
|  | 6 months prior | Community | 61 | 5.9 | 10 | 8 | 13 |
|  |  | Hospital | <10 | 0.2 | 38 | 18 | 80 |
|  | 12 months prior | Community | 82 | 5.9 | 14 | 11 | 17 |
|  |  | Hospital | 10 | 0.2 | 55 | 29 | 102 |
| **Nausea** | Post-COVID | Community | 41 | 5.7 | 7 | 5 | 10 |
|  |  | Hospital | <10 | 0.2 | 53 | 28 | 102 |
|  | 6 months prior | Community | 31 | 5.9 | 5 | 4 | 7 |
|  |  | Hospital | <10 | 0.2 | 22 | 8 | 58 |
|  | 12 months prior | Community | 27 | 5.9 | 5 | 3 | 7 |
|  |  | Hospital | <10 | 0.2 | 5 | 1 | 39 |
| **Anorexia** | Post-COVID | Community | 21 | 5.7 | 4 | 2 | 6 |
|  |  | Hospital | <10 | 0.2 | 12 | 3 | 47 |
|  | 6 months prior | Community | 19 | 5.9 | 3 | 2 | 5 |
|  |  | Hospital | <10 | 0.2 | 22 | 8 | 58 |
|  | 12 months prior | Community | 18 | 5.9 | 3 | 2 | 5 |
|  |  | Hospital | <10 | 0.2 | 5 | 1 | 39 |
| **Cognitive impairment** | Post-COVID | Community | 23 | 5.7 | 4 | 3 | 6 |
|  |  | Hospital | <10 | 0.2 | 18 | 6 | 55 |
|  | 6 months prior | Community | 18 | 5.9 | 3 | 2 | 5 |
|  |  | Hospital | <10 | 0.2 | 16 | 5 | 51 |
|  | 12 months prior | Community | 39 | 5.9 | 7 | 5 | 9 |
|  |  | Hospital | <10 | 0.2 | 27 | 11 | 66 |
| **Delirium** | Post-COVID | Community | 17 | 5.7 | 3 | 2 | 5 |
|  |  | Hospital | <10 | 0.2 | 24 | 9 | 63 |
|  | 6 months prior | Community | 18 | 5.9 | 3 | 2 | 5 |
|  |  | Hospital | <10 | 0.2 | 33 | 15 | 73 |
|  | 12 months prior | Community | 11 | 5.9 | 2 | 1 | 3 |
|  |  | Hospital | <10 | 0.2 | 27 | 11 | 65 |
| **Insomnia** | Post-COVID | Community | 79 | 5.7 | 14 | 11 | 17 |
|  |  | Hospital | <10 | 0.2 | 24 | 9 | 63 |
|  | 6 months prior | Community | 46 | 5.9 | 8 | 6 | 10 |
|  |  | Hospital | <10 | 0.2 | 44 | 22 | 87 |
|  | 12 months prior | Community | 61 | 5.9 | 10 | 8 | 13 |
|  |  | Hospital | <10 | 0.2 | 22 | 8 | 58 |
| **Dizziness** | Post-COVID | Community | 124 | 5.7 | 22 | 18 | 26 |
|  |  | Hospital | <10 | 0.2 | 54 | 28 | 103 |
|  | 6 months prior | Community | 124 | 5.9 | 21 | 18 | 25 |
|  |  | Hospital | 11 | 0.2 | 60 | 33 | 109 |
|  | 12 months prior | Community | 164 | 5.9 | 28 | 24 | 33 |
|  |  | Hospital | 10 | 0.2 | 55 | 29 | 102 |
| **Paresthesia** | Post-COVID | Community | 47 | 5.7 | 8 | 6 | 11 |
|  |  | Hospital | <10 | 0.2 | 12 | 3 | 47 |
|  | 6 months prior | Community | 50 | 5.9 | 9 | 6 | 11 |
|  |  | Hospital | <10 | 0.2 | 5 | 1 | 39 |
|  | 12 months prior | Community | 52 | 5.9 | 9 | 7 | 12 |
|  |  | Hospital | <10 | 0.2 | 27 | 11 | 65 |
| **Earache** | Post-COVID | Community | 82 | 5.7 | 14 | 12 | 18 |
|  |  | Hospital | <10 | 0.2 | 12 | 3 | 47 |
|  | 6 months prior | Community | 74 | 5.9 | 13 | 10 | 16 |
|  |  | Hospital | <10 | 0.2 | 27 | 11 | 65 |
|  | 12 months prior | Community | 82 | 5.9 | 14 | 11 | 17 |
|  |  | Hospital | <10 | 0.2 | 16 | 5 | 51 |
| **Sore throat** | Post-COVID | Community | 113 | 5.7 | 20 | 17 | 24 |
|  |  | Hospital | <10 | 0.2 | 18 | 6 | 55 |
|  | 6 months prior | Community | 123 | 5.9 | 21 | 18 | 25 |
|  |  | Hospital | <10 | 0.2 | 44 | 22 | 87 |
|  | 12 months prior | Community | 304 | 5.9 | 52 | 46 | 58 |
|  |  | Hospital | <10 | 0.2 | 22 | 8 | 58 |
| **Smell/taste loss** | Post-COVID | Community | 30 | 5.7 | 5 | 4 | 8 |
|  |  | Hospital | <10 | 0.2 | 12 | 3 | 47 |
|  | 6 months prior | Community | 30 | 5.9 | 5 | 4 | 7 |
|  |  | Hospital | <10 | 0.2 | 5 | 1 | 39 |
|  | 12 months prior* | Community | <10 | n/a | n/a | n/a | n/a |
|  |  | Hospital | <10 | n/a | n/a | n/a | n/a |
| **Tinnitus** | Post-COVID | Community | 36 | 5.7 | 6 | 5 | 9 |
|  |  | Hospital | <10 | 0.2 | 0 | n/a | n/a |
|  | 6 months prior | Community | 19 | 5.9 | 3 | 2 | 5 |
|  |  | Hospital | <10 | 0.2 | 11 | 3 | 44 |
|  | 12 months prior | Community | 26 | 5.9 | 4 | 3 | 6 |
|  |  | Hospital | <10 | 0.2 | 0 | n/a | n/a |
| **Skin rash** | Post-COVID | Community | 216 | 5.7 | 38 | 33 | 43 |
|  |  | Hospital | 12 | 0.2 | 71 | 40 | 125 |
|  | 6 months prior | Community | 188 | 5.9 | 32 | 28 | 37 |
|  |  | Hospital | <10 | 0.2 | 38 | 18 | 80 |
|  | 12 months prior | Community | 331 | 5.8 | 57 | 51 | 63 |
|  |  | Hospital | 13 | 0.2 | 71 | 41 | 123 |

*****No events for smell/taste loss using 12-month cohort.

### Table S3: Disease event rates (per 100,000 person-weeks).

| **Outcome** | **Time point** | **COVID exposure group** | **Number of events** | **Time (person-weeks (PW))** | **Rate (per 100,000 PW)** | **Lower CI** | **Upper CI** |
| --- | --- | --- | --- | --- | --- | --- | --- |
| **Asthma** | Post-COVID | Community | 322 | 5.3 | 60 | 54 | 67 |
|  |  | Hospital | 15 | 0.1 | 100 | 61 | 175 |
|  | 6 months prior | Community | 339 | 5.5 | 62 | 56 | 69 |
|  |  | Hospital | 14 | 0.2 | 87 | 52 | 155 |
|  | 12 months prior | Community | 358 | 5.5 | 65 | 59 | 72 |
|  |  | Hospital | 21 | 0.2 | 128 | 85 | 204 |
| **Lung fibrosis** | Post-COVID | Community | 16 | 5.7 | 3 | 2 | 5 |
|  |  | Hospital | <10 | 0.2 | 0 | n/a | n/a |
|  | 6 months prior | Community | <10 | 5.9 | 0 | n/a | n/a |
|  |  | Hospital | <10 | 0.2 | 5 | n/a | n/a |
|  | 12 months prior | Community | <10 | 5.9 | 1 | 0 | 3 |
|  |  | Hospital | <10 | 0.2 | 0 | n/a | n/a |
| **Heart failure** | Post-COVID | Community | 14 | 5.7 | 2 | 1 | 4 |
|  |  | Hospital | <10 | 0.2 | 54 | 29 | 116 |
|  | 6 months prior | Community | 16 | 5.9 | 3 | 2 | 5 |
|  |  | Hospital | <10 | 0.2 | 28 | 12 | 83 |
|  | 12 months prior | Community | 21 | 5.9 | 4 | 2 | 6 |
|  |  | Hospital | <10 | 0.2 | 28 | 12 | 84 |
| **IHD** | Post-COVID | Community | 48 | 5.7 | 8 | 6 | 11 |
|  |  | Hospital | 11 | 0.2 | 67 | 38 | 131 |
|  | 6 months prior | Community | 28 | 5.8 | 5 | 3 | 7 |
|  |  | Hospital | <10 | 0.2 | 22 | 8 | 81 |
|  | 12 months prior | Community | 59 | 5.8 | 10 | 8 | 13 |
|  |  | Hospital | <10 | 0.2 | 28 | 12 | 84 |
| **Hypertension** | Post-COVID | Community | 257 | 5.5 | 47 | 41 | 53 |
|  |  | Hospital | 36 | 0.1 | 244 | 178 | 344 |
|  | 6 months prior | Community | 138 | 5.6 | 24 | 21 | 29 |
|  |  | Hospital | 10 | 0.2 | 63 | 35 | 127 |
|  | 12 months prior | Community | 266 | 5.7 | 47 | 42 | 53 |
|  |  | Hospital | 34 | 0.2 | 211 | 153 | 302 |
| **Stroke** | Post-COVID | Community | 31 | 5.7 | 5 | 4 | 8 |
|  |  | Hospital | <10 | 0.2 | 37 | 17 | 96 |
|  | 6 months prior | Community | 20 | 5.9 | 3 | 2 | 6 |
|  |  | Hospital | <10 | 0.2 | 6 | n/a | n/a |
|  | 12 months prior | Community | 37 | 5.9 | 6 | 5 | 9 |
|  |  | Hospital | 12 | 0.2 | 67 | 39 | 127 |
| **PAD** | Post-COVID | Community | <10 | 5.7 | 0 | 0 | 4 |
|  |  | Hospital | <10 | 0.2 | 6 | n/a | n/a |
|  | 6 months prior | Community | <10 | 5.9 | 0 | 0 | 3 |
|  |  | Hospital | <10 | 0.2 | 5 | n/a | n/a |
|  | 12 months prior | Community | <10 | 5.9 | 1 | 1 | 3 |
|  |  | Hospital | <10 | 0.2 | 16 | 5 | 81 |
| **Anaemia** | Post-COVID | Community | 83 | 5.7 | 15 | 12 | 18 |
|  |  | Hospital | 14 | 0.2 | 87 | 52 | 155 |
|  | 6 months prior | Community | 49 | 5.8 | 8 | 6 | 11 |
|  |  | Hospital | <10 | 0.2 | 46 | 23 | 102 |
|  | 12 months prior | Community | 108 | 5.8 | 19 | 15 | 23 |
|  |  | Hospital | 19 | 0.2 | 107 | 69 | 175 |
| **VTE** | Post-COVID | Community | 56 | 5.7 | 10 | 8 | 13 |
|  |  | Hospital | 30 | 0.2 | 185 | 130 | 271 |
|  | 6 months prior | Community | 37 | 5.9 | 6 | 5 | 9 |
|  |  | Hospital | <10 | 0.2 | 22 | 8 | 80 |
|  | 12 months prior | Community | 28 | 5.9 | 5 | 3 | 7 |
|  |  | Hospital | <10 | 0.2 | 11 | 2 | 111 |
| **Renal failure** | Post-COVID | Community | 62 | 5.7 | 11 | 9 | 14 |
|  |  | Hospital | 24 | 0.2 | 149 | 101 | 230 |
|  | 6 months prior | Community | 33 | 5.8 | 6 | 4 | 8 |
|  |  | Hospital | <10 | 0.2 | 23 | 9 | 81 |
|  | 12 months prior | Community | 63 | 5.8 | 11 | 8 | 14 |
|  |  | Hospital | <10 | 0.2 | 51 | 27 | 109 |
| **GORD** | Post-COVID | Community | 131 | 5.6 | 23 | 20 | 28 |
|  |  | Hospital | <10 | 0.2 | 18 | 6 | 88 |
|  | 6 months prior | Community | 120 | 5.8 | 21 | 17 | 25 |
|  |  | Hospital | <10 | 0.2 | 39 | 19 | 93 |
|  | 12 months prior | Community | 128 | 5.8 | 22 | 19 | 26 |
|  |  | Hospital | <10 | 0.2 | 22 | 8 | 80 |
| **Liver disease** | Post-COVID | Community | 77 | 5.7 | 14 | 11 | 17 |
|  |  | Hospital | <10 | 0.2 | 30 | 13 | 90 |
|  | 6 months prior | Community | 39 | 5.8 | 7 | 5 | 9 |
|  |  | Hospital | <10 | 0.2 | 17 | 5 | 82 |
|  | 12 months prior | Community | 69 | 5.8 | 12 | 9 | 15 |
|  |  | Hospital | <10 | 0.2 | 44 | 23 | 100 |
| **Diabetes** | Post-COVID | Community | 198 | 5.4 | 36 | 32 | 42 |
|  |  | Hospital | 42 | 0.1 | 303 | 225 | 416 |
|  | 6 months prior | Community | 82 | 5.6 | 15 | 12 | 18 |
|  |  | Hospital | <10 | 0.1 | 61 | 32 | 128 |
|  | 12 months prior | Community | 205 | 5.6 | 37 | 32 | 42 |
|  |  | Hospital | 19 | 0.1 | 128 | 82 | 208 |
| **Adrenal disease** | Post-COVID | Community | <10 | 5.7 | 0 | n/a | n/a |
|  |  | Hospital | <10 | 0.2 | 6 | n/a | n/a |
|  | 6 months prior | Community | <10 | 5.9 | 0 | n/a | n/a |
|  |  | Hospital | <10 | 0.2 | 0 | n/a | n/a |
|  | 12 months prior | Community | <10 | 5.9 | 0 | n/a | n/a |
|  |  | Hospital | <10 | 0.2 | 0 | n/a | n/a |
| **Arthritis** | Post-COVID | Community | 93 | 5.6 | 17 | 14 | 20 |
|  |  | Hospital | <10 | 0.2 | 51 | 26 | 113 |
|  | 6 months prior | Community | 59 | 5.8 | 10 | 8 | 13 |
|  |  | Hospital | <10 | 0.2 | 53 | 28 | 112 |
|  | 12 months prior | Community | 162 | 5.8 | 28 | 24 | 33 |
|  |  | Hospital | 13 | 0.2 | 76 | 45 | 140 |
| **Anxiety** | Post-COVID | Community | 502 | 5.4 | 93 | 86 | 102 |
|  |  | Hospital | 16 | 0.2 | 100 | 62 | 171 |
|  | 6 months prior | Community | 347 | 5.5 | 63 | 57 | 70 |
|  |  | Hospital | 13 | 0.2 | 74 | 44 | 136 |
|  | 12 months prior | Community | 495 | 5.5 | 90 | 82 | 98 |
|  |  | Hospital | 12 | 0.2 | 69 | 40 | 130 |
| **Depression** | Post-COVID | Community | 346 | 5.5 | 63 | 57 | 70 |
|  |  | Hospital | <10 | 0.2 | 55 | 29 | 117 |
|  | 6 months prior | Community | 223 | 5.6 | 40 | 35 | 45 |
|  |  | Hospital | <10 | 0.2 | 46 | 23 | 102 |
|  | 12 months prior | Community | 358 | 5.6 | 63 | 57 | 71 |
|  |  | Hospital | 10 | 0.2 | 57 | 31 | 116 |

### Table S4: Prescription event rates (per 100,000 person-weeks).

| **Outcome** | **Time point** | **COVID exposure group** | **Number of events** | **Time (person-weeks (PW))** | **Rate (per 100,000 PW)** | **Lower CI** | **Upper CI** |
| --- | --- | --- | --- | --- | --- | --- | --- |
| **Diuretics** | Post-COVID | Community | 22 | 5.6 | 4 | 3 | 6 |
|  |  | Hospital | <10 | 0.2 | 32 | 13 | 76 |
|  | 6 months prior | Community | 17 | 5.8 | 3 | 2 | 5 |
|  |  | Hospital | <10 | 0.2 | 12 | 3 | 47 |
|  | 12 months prior | Community | 32 | 5.8 | 6 | 4 | 8 |
|  |  | Hospital | <10 | 0.2 | 35 | 16 | 79 |
| **Bronchodilators** | Post-COVID | Community | 314 | 5.1 | 61 | 55 | 68 |
|  |  | Hospital | 28 | 0.1 | 212 | 147 | 308 |
|  | 6 months prior | Community | 229 | 5.3 | 43 | 38 | 49 |
|  |  | Hospital | 11 | 0.1 | 77 | 43 | 139 |
|  | 12 months prior | Community | 315 | 5.3 | 59 | 53 | 66 |
|  |  | Hospital | 13 | 0.1 | 90 | 52 | 155 |
| **ICS** | Post-COVID | Community | 173 | 5.3 | 33 | 28 | 38 |
|  |  | Hospital | 20 | 0.1 | 144 | 93 | 224 |
|  | 6 months prior | Community | 148 | 5.4 | 27 | 23 | 32 |
|  |  | Hospital | 10 | 0.2 | 66 | 35 | 122 |
|  | 12 months prior | Community | 173 | 5.5 | 32 | 27 | 37 |
|  |  | Hospital | 10 | 0.2 | 65 | 35 | 121 |
| **Paracetamol** | Post-COVID | Community | 104 | 5.5 | 19 | 16 | 23 |
|  |  | Hospital | 43 | 0.1 | 304 | 226 | 411 |
|  | 6 months prior | Community | 109 | 5.7 | 19 | 16 | 23 |
|  |  | Hospital | 16 | 0.2 | 103 | 63 | 168 |
|  | 12 months prior | Community | 84 | 5.7 | 15 | 12 | 18 |
|  |  | Hospital | <10 | 0.2 | 45 | 21 | 94 |
| **NSAIDs** | Post-COVID | Community | 428 | 5.1 | 84 | 76 | 92 |
|  |  | Hospital | 39 | 0.1 | 303 | 221 | 414 |
|  | 6 months prior | Community | 443 | 5.2 | 85 | 77 | 93 |
|  |  | Hospital | 22 | 0.1 | 158 | 104 | 240 |
|  | 12 months prior | Community | 475 | 5.2 | 91 | 83 | 99 |
|  |  | Hospital | 22 | 0.1 | 158 | 104 | 240 |
| **Opioids** | Post-COVID | Community | 418 | 5.1 | 82 | 74 | 90 |
|  |  | Hospital | 44 | 0.1 | 378 | 282 | 508 |
|  | 6 months prior | Community | 401 | 5.2 | 77 | 69 | 84 |
|  |  | Hospital | 40 | 0.1 | 319 | 234 | 435 |
|  | 12 months prior | Community | 432 | 5.2 | 83 | 75 | 91 |
|  |  | Hospital | 29 | 0.1 | 231 | 161 | 333 |
| **Neuropathic pain medication** | Post-COVID | Community | 247 | 5.4 | 46 | 41 | 52 |
|  |  | Hospital | 20 | 0.1 | 143 | 92 | 222 |
|  | 6 months prior | Community | 189 | 5.5 | 34 | 30 | 39 |
|  |  | Hospital | 15 | 0.2 | 99 | 59 | 164 |
|  | 12 months prior | Community | 228 | 5.5 | 41 | 36 | 47 |
|  |  | Hospital | 19 | 0.2 | 124 | 79 | 194 |

### Table S5: Health care utilization (HCU) event rates (per 100,000 person-weeks).

| **Outcome** | **Time point** | **COVID exposure group** | **Number of events** | **Time (person-weeks (PW))** | **Rate (per 100,000 PW)** | **Lower CI** | **Upper CI** |
| --- | --- | --- | --- | --- | --- | --- | --- |
| **HCU** | Post-COVID | Community | 95700 | 4.9 | 19405 | 19142 | 19673 |
|  |  | Hospital | 7571 | 0.1 | 52775 | 50570 | 55105 |
|  | 6 months prior | Community | 57007 | 5.2 | 10862 | 10678 | 11050 |
|  |  | Hospital | 4516 | 0.2 | 28841 | 27075 | 30753 |
|  | 12 months prior | Community | 76022 | 5.0 | 15100 | 14886 | 15318 |
|  |  | Hospital | 5106 | 0.2 | 32728 | 30990 | 34590 |

### Table S6: Hazard ratios (95% CI) for differences in outcome event rates between patients with community COVID-19 and patients hospitalised for COVID-19 in the post-COVID-19 population

| **Type of outcome** | **Outcome** | **HR** | **HR 95% CI** | **p-value** |
| --- | --- | --- | --- | --- |
| **Symptoms** | General pain | 1.48 | 0.36-6.10 | 0.587 |
|  | Chest pain | 3.2 | 2.14-5.78 | <0.001 |
|  | Abdominal pain | 1.63 | 1.01-2.61 | 0.044 |
|  | Headache | 1.35 | 0.74-2.47 | 0.327 |
|  | Joint pain | 1.78 | 1.33-2.39 | <0.001 |
|  | Muscle pain | 2.18 | 0.68-7.01 | 0.190 |
|  | Neuropathic pain | 4.27 | 0.53-34.11 | 0.171 |
|  | Fatigue | 2.34 | 1.43-3.82 | 0.001 |
|  | Fever | 6.47 | 2.49-16.80 | <0.001 |
|  | Breathlessness | 6.33 | 5.02-7.98 | <0.001 |
|  | Cough | 3.06 | 2.01-4.64 | <0.001 |
|  | Chest tightness | n/a | n/a | n/a |
|  | Palpitations | 0.99 | 0.31-3.12 | 0.987 |
|  | Diarrhoea | 3.23 | 1.40-7.45 | 0.006 |
|  | Nausea | 7.52 | 3.65-15.47 | <0.001 |
|  | Anorexia | 3.25 | 0.76-13.86 | 0.111 |
|  | Cognitive impairment | 4.47 | 1.34-14.88 | 0.015 |
|  | Delirium | 8.09 | 2.72-24.05 | <0.001 |
|  | Insomnia | 1.73 | 0.63-4.72 | 0.285 |
|  | Dizziness | 2.49 | 1.27-4.90 | 0.008 |
|  | Paraesthesia | 1.45 | 0.35-5.98 | 0.606 |
|  | Earache | 0.83 | 0.20-3.38 | 0.796 |
|  | Sore throat | 0.9 | 0.29-2.85 | 0.864 |
|  | Smell or taste loss | 2.27 | 0.54-9.50 | 0.262 |
|  | Tinnitus | n/a | n/a | n/a |
|  | Skin rash | 1.9 | 1.06-3.40 | 0.030 |
| **Diseases** | Asthma | 1.65 | 0.98-2.77 | 0.057 |
|  | Lung fibrosis | n/a | n/a | n/a |
|  | Heart failure | 22.35 | 9.67-51.64 | <0.001 |
|  | IHD | 7.9 | 4.1-15.22 | <0.001 |
|  | Hypertension | 5.26 | 3.71-7.45 | <0.001 |
|  | Stroke | 6.68 | 2.79-16.02 | <0.001 |
|  | PAD | 16.68 | 1.51-183.96 | 0.022 |
|  | Anaemia | 5.93 | 3.37-10.45 | <0.001 |
|  | VTE | 18.5 | 11.87-28.83 | <0.001 |
|  | Renal failure | 13.71 | 8.56-21.97 | <0.001 |
|  | GORD | 0.77 | 0.25-2.42 | 0.656 |
|  | Liver disease | 2.23 | 0.90-5.50 | 0.083 |
|  | Diabetes | 8.34 | 5.98-11.63 | <0.001 |
|  | Adrenal disease | 32.63 | 2.04-521.78 | 0.014 |
|  | Arthritis | 3.07 | 1.49-6.32 | 0.002 |
|  | Anxiety | 1.06 | 0.65-1.75 | 0.811 |
|  | Depression | 0.88 | 0.45-1.70 | 0.694 |
| **Medications** | Diuretics | 8.20 | 3.11-21.66 | <0.001 |
|  | Bronchodilators | 3.51 | 2.38-5.16 | <0.001 |
|  | ICS | 4.45 | 2.80-7.07 | <0.001 |
|  | Paracetamol | 16.38 | 11.48-23.37 | <0.001 |
|  | NSAIDs | 3.68 | 2.65-5.11 | <0.001 |
|  | Opiates | 4.69 | 3.44-6.40 | <0.001 |
|  | Neuropathic pain medication | 3.17 | 2.01-5.00 | <0.001 |

### Table S7: Symptom event rates (per 100,000 person-weeks) in men over 50 years old with community COVID-19.

| **Outcome** | **Time point** | **Number of events** | **Time (person-weeks (PW))** | **Rate (per 100,000 PW)** | **Lower CI** | **Upper CI** |
| --- | --- | --- | --- | --- | --- | --- |
| **General pain** | Post-COVID | <10 | 0.7 | 6 | 2 | 15 |
|  | 6 months prior | <10 | 0.7 | 12 | 6 | 23 |
|  | 12 months prior | <10 | 0.7 | 11 | 5 | 21 |
| **Chest pain** | Post-COVID | 47 | 0.7 | 65 | 49 | 87 |
|  | 6 months prior | 28 | 0.7 | 38 | 26 | 55 |
|  | 12 months prior | 41 | 0.7 | 55 | 41 | 75 |
| **Abdominal pain** | Post-COVID | 39 | 0.7 | 54 | 40 | 74 |
|  | 6 months prior | 25 | 0.7 | 34 | 23 | 50 |
|  | 12 months prior | 36 | 0.7 | 49 | 35 | 67 |
| **Headache** | Post-COVID | 14 | 0.7 | 19 | 12 | 33 |
|  | 6 months prior | 11 | 0.7 | 15 | 8 | 27 |
|  | 12 months prior | 21 | 0.7 | 28 | 18 | 43 |
| **Joint pain** | Post-COVID | 137 | 0.7 | 195 | 165 | 230 |
|  | 6 months prior | 139 | 0.7 | 190 | 161 | 225 |
|  | 12 months prior | 234 | 0.7 | 326 | 287 | 371 |
| **Muscle pain** | Post-COVID | <10 | 0.7 | 11 | 6 | 22 |
|  | 6 months prior | <10 | 0.7 | 8 | 4 | 18 |
|  | 12 months prior | 10 | 0.7 | 13 | 7 | 25 |
| **Neuropathic pain** | Post-COVID | <10 | 0.7 | 1 | 0 | 10 |
|  | 6 months prior | <10 | 0.7 | 1 | 0 | 10 |
|  | 12 months prior | <10 | 0.7 | 1 | 0 | 10 |
| **Fatigue** | Post-COVID | 31 | 0.7 | 43 | 30 | 61 |
|  | 6 months prior | 12 | 0.7 | 16 | 9 | 28 |
|  | 12 months prior | 15 | 0.7 | 20 | 12 | 33 |
| **Fever** | Post-COVID | <10 | 0.7 | 4 | 1 | 13 |
|  | 6 months prior | <10 | 0.7 | 5 | 2 | 14 |
|  | 12 months prior | <10 | 0.7 | 5 | 2 | 14 |
| **Breathlessness** | Post-COVID | 115 | 0.7 | 162 | 135 | 195 |
|  | 6 months prior | 63 | 0.7 | 85 | 67 | 109 |
|  | 12 months prior | 68 | 0.7 | 92 | 72 | 117 |
| **Cough** | Post-COVID | 51 | 0.7 | 72 | 55 | 95 |
|  | 6 months prior | 57 | 0.7 | 77 | 60 | 100 |
|  | 12 months prior | 129 | 0.7 | 175 | 147 | 208 |
| **Chest tightness** | Post-COVID | <10 | 0.7 | 3 | 1 | 11 |
|  | 6 months prior | <10 | 0.7 | 3 | 1 | 11 |
|  | 12 months prior | <10 | 0.7 | 5 | 2 | 14 |
| **Palpitations** | Post-COVID | 10 | 0.7 | 14 | 7 | 26 |
|  | 6 months prior | <10 | 0.7 | 7 | 3 | 16 |
|  | 12 months prior | <10 | 0.7 | 11 | 5 | 21 |
| **Diarrhoea** | Post-COVID | 11 | 0.7 | 15 | 8 | 27 |
|  | 6 months prior | 12 | 0.7 | 16 | 9 | 28 |
|  | 12 months prior | 13 | 0.7 | 17 | 10 | 30 |
| **Nausea** | Post-COVID | <10 | 0.7 | 7 | 3 | 17 |
|  | 6 months prior | <10 | 0.7 | 4 | 1 | 12 |
|  | 12 months prior | <10 | 0.7 | 3 | 1 | 11 |
| **Anorexia** | Post-COVID | <10 | 0.7 | 7 | 3 | 17 |
|  | 6 months prior | <10 | 0.7 | 7 | 3 | 16 |
|  | 12 months prior | <10 | 0.7 | 3 | 1 | 11 |
| **Cognitive impairment** | Post-COVID | <10 | 0.7 | 7 | 3 | 17 |
|  | 6 months prior | <10 | 0.7 | 8 | 4 | 18 |
|  | 12 months prior | 10 | 0.7 | 13 | 7 | 25 |
| **Delirium** | Post-COVID | <10 | 0.7 | 7 | 3 | 17 |
|  | 6 months prior | <10 | 0.7 | 1 | 0 | 10 |
|  | 12 months prior | <10 | 0.7 | 4 | 1 | 12 |
| **Insomnia** | Post-COVID | <10 | 0.7 | 12 | 6 | 24 |
|  | 6 months prior | <10 | 0.7 | 9 | 4 | 20 |
|  | 12 months prior | <10 | 0.7 | 5 | 2 | 14 |
| **Dizziness** | Post-COVID | 25 | 0.7 | 35 | 23 | 51 |
|  | 6 months prior | 20 | 0.7 | 27 | 17 | 42 |
|  | 12 months prior | 32 | 0.7 | 43 | 30 | 61 |
| **Paraesthesia** | Post-COVID | 10 | 0.7 | 14 | 7 | 26 |
|  | 6 months prior | 13 | 0.7 | 17 | 10 | 30 |
|  | 12 months prior | 11 | 0.7 | 15 | 8 | 27 |
| **Earache** | Post-COVID | <10 | 0.7 | 6 | 2 | 15 |
|  | 6 months prior | 15 | 0.7 | 20 | 12 | 33 |
|  | 12 months prior | <10 | 0.7 | 4 | 1 | 12 |
| **Sore throat** | Post-COVID | <10 | 0.7 | 8 | 4 | 19 |
|  | 6 months prior | <10 | 0.7 | 12 | 6 | 23 |
|  | 12 months prior | 15 | 0.7 | 20 | 12 | 33 |
| **Smell/taste loss** | Post-COVID | <10 | 0.7 | 4 | 1 | 13 |
|  | 6 months prior | <10 | 0.7 | 3 | 1 | 11 |
| **Tinnitus** | Post-COVID | <10 | 0.7 | 8 | 4 | 18 |
|  | 6 months prior | <10 | 0.7 | 4 | 1 | 12 |
|  | 12 months prior | <10 | 0.7 | 4 | 1 | 12 |
| **Skin rash** | Post-COVID | 19 | 0.7 | 26 | 17 | 41 |
|  | 6 months prior | 18 | 0.7 | 24 | 15 | 38 |
|  | 12 months prior | 35 | 0.7 | 47 | 34 | 66 |

### Table S8: Disease outcome events (per 100,000 person-weeks) in men over 50 years old with community COVID-19.

| **Outcome** | **Time point** | **Number of events** | **Time (person-weeks (PW))** | **Rate (per 100,000 PW)** | **Lower CI** | **Upper CI** |
| --- | --- | --- | --- | --- | --- | --- |
| **Asthma** | Post-COVID | 42 | 0.7 | 62 | 46 | 84 |
|  | 6 months prior | 36 | 0.7 | 52 | 38 | 72 |
|  | 12 months prior | 48 | 0.7 | 69 | 52 | 92 |
| **Lung fibrosis** | Post-COVID | 10 | 0.7 | 14 | 7 | 26 |
|  | 6 months prior | <10 | 0.7 | 0 | n/a | n/a |
|  | 12 months prior | <10 | 0.7 | 1 | 0 | 10 |
| **Heart failure** | Post-COVID | <10 | 0.7 | 13 | 7 | 24 |
|  | 6 months prior | <10 | 0.7 | 11 | 5 | 22 |
|  | 12 months prior | 12 | 0.7 | 16 | 9 | 29 |
| **IHD** | Post-COVID | 29 | 0.7 | 41 | 29 | 59 |
|  | 6 months prior | 16 | 0.7 | 22 | 14 | 36 |
|  | 12 months prior | 39 | 0.7 | 54 | 39 | 74 |
| **Hypertension** | Post-COVID | 95 | 0.6 | 147 | 120 | 179 |
|  | 6 months prior | 51 | 0.7 | 78 | 59 | 103 |
|  | 12 months prior | 99 | 0.7 | 152 | 124 | 185 |
| **Stroke** | Post-COVID | 17 | 0.7 | 24 | 15 | 38 |
|  | 6 months prior | 10 | 0.7 | 14 | 7 | 25 |
|  | 12 months prior | 17 | 0.7 | 23 | 14 | 37 |
| **PAD** | Post-COVID | <10 | 0.7 | 3 | 1 | 11 |
|  | 6 months prior | <10 | 0.7 | 1 | 0 | 10 |
|  | 12 months prior | <10 | 0.7 | 8 | 4 | 18 |
| **Anaemia** | Post-COVID | 16 | 0.7 | 22 | 14 | 37 |
|  | 6 months prior | <10 | 0.7 | 12 | 6 | 24 |
|  | 12 months prior | 20 | 0.7 | 27 | 18 | 42 |
| **VTE** | Post-COVID | 13 | 0.7 | 18 | 11 | 31 |
|  | 6 months prior | 14 | 0.7 | 19 | 11 | 32 |
|  | 12 months prior | <10 | 0.7 | 8 | 4 | 18 |
| **Renal failure** | Post-COVID | 17 | 0.7 | 24 | 15 | 38 |
|  | 6 months prior | 10 | 0.7 | 14 | 7 | 25 |
|  | 12 months prior | 21 | 0.7 | 29 | 19 | 44 |
| **GORD** | Post-COVID | 25 | 0.7 | 35 | 24 | 52 |
|  | 6 months prior | 23 | 0.7 | 31 | 21 | 47 |
|  | 12 months prior | 16 | 0.7 | 22 | 13 | 36 |
| **Liver disease** | Post-COVID | 19 | 0.7 | 27 | 17 | 42 |
|  | 6 months prior | 11 | 0.7 | 15 | 8 | 27 |
|  | 12 months prior | 15 | 0.7 | 20 | 12 | 34 |
| **Diabetes** | Post-COVID | 63 | 0.6 | 101 | 79 | 129 |
|  | 6 months prior | 26 | 0.6 | 41 | 28 | 60 |
|  | 12 months prior | 64 | 0.6 | 101 | 79 | 129 |
| **Adrenal disease** | Post-COVID | <10 | 0.7 | 0 | n/a | n/a |
|  | 6 months prior | <10 | 0.7 | 0 | n/a | n/a |
|  | 12 months prior | <10 | 0.7 | 1 | 0 | 10 |
| **Arthritis** | Post-COVID | 31 | 0.7 | 44 | 31 | 63 |
|  | 6 months prior | 21 | 0.7 | 29 | 19 | 45 |
|  | 12 months prior | 54 | 0.7 | 76 | 58 | 99 |
| **Anxiety** | Post-COVID | 24 | 0.7 | 34 | 23 | 51 |
|  | 6 months prior | 26 | 0.7 | 36 | 24 | 53 |
|  | 12 months prior | 19 | 0.7 | 26 | 17 | 41 |
| **Depression** | Post-COVID | 27 | 0.7 | 38 | 26 | 56 |
|  | 6 months prior | 11 | 0.7 | 15 | 8 | 27 |
|  | 12 months prior | 23 | 0.7 | 32 | 21 | 48 |

### Table S9: Prescription event rates per (100,000 person-weeks) in men over 50 years old with community COVID-19.

| **Outcome** | **Time point** | **Number of events** | **Time (person-weeks (PW))** | **Rate (per 100,000 PW)** | **Lower CI** | **Upper CI** |
| --- | --- | --- | --- | --- | --- | --- |
| **Diuretics** | Post-COVID | <10 | 0.7 | 12 | 6 | 24 |
|  | 6 months prior | <10 | 0.7 | 9 | 4 | 19 |
|  | 12 months prior | 12 | 0.7 | 17 | 10 | 30 |
| **Bronchodilators** | Post-COVID | 47 | 0.6 | 74 | 56 | 99 |
|  | 6 months prior | 21 | 0.7 | 32 | 21 | 49 |
|  | 12 months prior | 42 | 0.7 | 64 | 47 | 86 |
| **ICS** | Post-COVID | 23 | 0.6 | 36 | 24 | 54 |
|  | 6 months prior | 16 | 0.7 | 24 | 15 | 39 |
|  | 12 months prior | 16 | 0.7 | 24 | 15 | 39 |
| **Paracetamol** | Post-COVID | 39 | 0.7 | 58 | 43 | 80 |
|  | 6 months prior | 34 | 0.7 | 49 | 35 | 69 |
|  | 12 months prior | 18 | 0.7 | 26 | 16 | 41 |
| **NSAID** | Post-COVID | 60 | 0.6 | 104 | 81 | 134 |
|  | 6 months prior | 64 | 0.6 | 108 | 85 | 138 |
|  | 12 months prior | 83 | 0.6 | 141 | 114 | 175 |
| **Opioid medication** | Post-COVID | 80 | 0.6 | 134 | 108 | 167 |
|  | 6 months prior | 80 | 0.6 | 130 | 105 | 162 |
|  | 12 months prior | 77 | 0.6 | 126 | 101 | 157 |
| **Neuropathic pain medication** | Post-COVID | 36 | 0.7 | 54 | 39 | 75 |
|  | 6 months prior | 29 | 0.7 | 43 | 30 | 61 |
|  | 12 months prior | 36 | 0.7 | 53 | 38 | 73 |

### Table S10: Symptom event rates (per 100,000 person-weeks) in men 50 years old or younger with community COVID-19.

| **Outcome** | **Time point** | **Number of events** | **Time (person-weeks (PW))** | **Rate (per 100,000 PW)** | **Lower CI** | **Upper CI** |
| --- | --- | --- | --- | --- | --- | --- |
| **General pain** | Post-COVID | <10 | 1.9 | 5 | 3 | 9 |
|  | 6 months prior | <10 | 1.9 | 3 | 1 | 7 |
|  | 12 months prior | <10 | 1.9 | 3 | 1 | 7 |
| **Chest pain** | Post-COVID | 69 | 1.9 | 37 | 29 | 47 |
|  | 6 months prior | 39 | 1.9 | 20 | 15 | 28 |
|  | 12 months prior | 58 | 1.9 | 30 | 24 | 39 |
| **Abdominal pain** | Post-COVID | 69 | 1.9 | 37 | 29 | 47 |
|  | 6 months prior | 44 | 1.9 | 23 | 17 | 31 |
|  | 12 months prior | 63 | 1.9 | 33 | 26 | 42 |
| **Headache** | Post-COVID | 49 | 1.9 | 26 | 20 | 35 |
|  | 6 months prior | 35 | 1.9 | 18 | 13 | 26 |
|  | 12 months prior | 63 | 1.9 | 33 | 26 | 42 |
| **Joint pain** | Post-COVID | 187 | 1.8 | 102 | 88 | 117 |
|  | 6 months prior | 182 | 1.9 | 96 | 83 | 111 |
|  | 12 months prior | 258 | 1.9 | 137 | 122 | 155 |
| **Muscle pain** | Post-COVID | 10 | 1.9 | 5 | 3 | 10 |
|  | 6 months prior | <10 | 1.9 | 2 | 1 | 6 |
|  | 12 months prior | 13 | 1.9 | 7 | 4 | 12 |
| **Neuropathic pain** | Post-COVID | <10 | 1.9 | 1 | 0 | 4 |
|  | 6 months prior | <10 | 1.9 | 1 | 0 | 4 |
|  | 12 months prior | <10 | 1.9 | 1 | 0 | 4 |
| **Fatigue** | Post-COVID | 33 | 1.9 | 18 | 13 | 25 |
|  | 6 months prior | 14 | 1.9 | 7 | 4 | 12 |
|  | 12 months prior | 30 | 1.9 | 16 | 11 | 22 |
| **Fever** | Post-COVID | <10 | 1.9 | 3 | 1 | 7 |
|  | 6 months prior | 11 | 1.9 | 6 | 3 | 10 |
|  | 12 months prior | <10 | 1.9 | 3 | 1 | 7 |
| **Breathlessness** | Post-COVID | 74 | 1.9 | 40 | 32 | 50 |
|  | 6 months prior | 37 | 1.9 | 19 | 14 | 27 |
|  | 12 months prior | 20 | 1.9 | 10 | 7 | 16 |
| **Cough** | Post-COVID | 39 | 1.9 | 21 | 15 | 29 |
|  | 6 months prior | 35 | 1.9 | 18 | 13 | 26 |
|  | 12 months prior | 108 | 1.9 | 57 | 47 | 69 |
| **Chest tightness** | Post-COVID | <10 | 1.9 | 3 | 1 | 6 |
|  | 6 months prior | <10 | 1.9 | 3 | 1 | 7 |
|  | 12 months prior | <10 | 1.9 | 0 | n/a | n/a |
| **Palpitations** | Post-COVID | 23 | 1.9 | 12 | 8 | 19 |
|  | 6 months prior | <10 | 1.9 | 5 | 2 | 9 |
|  | 12 months prior | 12 | 1.9 | 6 | 4 | 11 |
| **Diarrhoea** | Post-COVID | 14 | 1.9 | 8 | 4 | 13 |
|  | 6 months prior | <10 | 1.9 | 5 | 2 | 9 |
|  | 12 months prior | 21 | 1.9 | 11 | 7 | 17 |
| **Nausea** | Post-COVID | <10 | 1.9 | 3 | 1 | 6 |
|  | 6 months prior | <10 | 1.9 | 2 | 1 | 5 |
|  | 12 months prior | <10 | 1.9 | 1 | 0 | 4 |
| **Anorexia** | Post-COVID | <10 | 1.9 | 1 | 0 | 4 |
|  | 6 months prior | <10 | 1.9 | 1 | 0 | 4 |
|  | 12 months prior | <10 | 1.9 | 1 | 0 | 4 |
| **Cognitive impairment** | Post-COVID | <10 | 1.9 | 1 | 0 | 4 |
|  | 6 months prior | <10 | 1.9 | 0 | n/a | n/a |
|  | 12 months prior | <10 | 1.9 | 1 | 0 | 4 |
| **Delirium** | Post-COVID | <10 | 1.9 | 1 | 0 | 4 |
|  | 6 months prior | <10 | 1.9 | 1 | 0 | 4 |
|  | 12 months prior | <10 | 1.9 | 1 | 0 | 4 |
| **Insomnia** | Post-COVID | 19 | 1.9 | 10 | 7 | 16 |
|  | 6 months prior | <10 | 1.9 | 3 | 1 | 7 |
|  | 12 months prior | 17 | 1.9 | 9 | 6 | 14 |
| **Dizziness** | Post-COVID | 13 | 1.9 | 7 | 4 | 12 |
|  | 6 months prior | 18 | 1.9 | 9 | 6 | 15 |
|  | 12 months prior | 19 | 1.9 | 10 | 6 | 16 |
| **Paranesthesia** | Post-COVID | <10 | 1.9 | 3 | 1 | 7 |
|  | 6 months prior | <10 | 1.9 | 4 | 2 | 8 |
|  | 12 months prior | <10 | 1.9 | 3 | 1 | 7 |
| **Earache** | Post-COVID | 12 | 1.9 | 6 | 4 | 11 |
|  | 6 months prior | 16 | 1.9 | 8 | 5 | 14 |
|  | 12 months prior | 17 | 1.9 | 9 | 6 | 14 |
| **Sore throat** | Post-COVID | 34 | 1.9 | 18 | 13 | 26 |
|  | 6 months prior | 30 | 1.9 | 16 | 11 | 22 |
|  | 12 months prior | 90 | 1.9 | 47 | 38 | 58 |
| **Smell/taste loss** | Post-COVID | <10 | 1.9 | 3 | 1 | 7 |
|  | 6 months prior | <10 | 1.9 | 3 | 1 | 7 |
| **Tinnitus** | Post-COVID | <10 | 1.9 | 5 | 3 | 9 |
|  | 6 months prior | 10 | 1.9 | 5 | 3 | 10 |
|  | 12 months prior | <10 | 1.9 | 4 | 2 | 8 |
| **Skin rash** | Post-COVID | 47 | 1.9 | 25 | 19 | 34 |
|  | 6 months prior | 43 | 1.9 | 23 | 17 | 30 |
|  | 12 months prior | 87 | 1.9 | 46 | 37 | 56 |

### Table S11: Disease outcome events (per 100,000 person-weeks) in men 50 years old or younger with community COVID-19.

| **Outcome** | **Time point** | **Number of events** | **Time (person-weeks (PW))** | **Rate (per 100,000 PW)** | **Lower CI** | **Upper CI** |
| --- | --- | --- | --- | --- | --- | --- |
| **Asthma** | Post-COVID | 94 | 1.8 | 53 | 43 | 65 |
|  | 6 months prior | 106 | 1.8 | 59 | 48 | 71 |
|  | 12 months prior | 106 | 1.8 | 58 | 48 | 70 |
| **Lung fibrosis** | Post-COVID | <10 | 1.9 | 1 | 0 | 4 |
|  | 6 months prior | <10 | 1.9 | 0 | n/a | n/a |
|  | 12 months prior | <10 | 1.9 | 0 | n/a | n/a |
| **Heart failure** | Post-COVID | <10 | 1.9 | 1 | 0 | 4 |
|  | 6 months prior | <10 | 1.9 | 0 | n/a | n/a |
|  | 12 months prior | <10 | 1.9 | 1 | 0 | 4 |
| **IHD** | Post-COVID | <10 | 1.9 | 1 | 0 | 4 |
|  | 6 months prior | <10 | 1.9 | 1 | 0 | 4 |
|  | 12 months prior | <10 | 1.9 | 1 | 0 | 4 |
| **Hypertension** | Post-COVID | 24 | 1.8 | 13 | 9 | 19 |
|  | 6 months prior | 14 | 1.9 | 7 | 4 | 13 |
|  | 12 months prior | 36 | 1.9 | 19 | 14 | 26 |
| **Stroke** | Post-COVID | <10 | 1.9 | 1 | 0 | 4 |
|  | 6 months prior | <10 | 1.9 | 1 | 0 | 4 |
|  | 12 months prior | <10 | 1.9 | 1 | 0 | 4 |
| **PAD** | Post-COVID | <10 | 1.9 | 0 | n/a | n/a |
|  | 6 months prior | <10 | 1.9 | 0 | n/a | n/a |
|  | 12 months prior | <10 | 1.9 | 0 | n/a | n/a |
| **Anaemia** | Post-COVID | <10 | 1.9 | 1 | 0 | 4 |
|  | 6 months prior | <10 | 1.9 | 1 | 0 | 4 |
|  | 12 months prior | <10 | 1.9 | 1 | 0 | 4 |
| **VTE** | Post-COVID | <10 | 1.9 | 5 | 3 | 9 |
|  | 6 months prior | <10 | 1.9 | 3 | 1 | 7 |
|  | 12 months prior | <10 | 1.9 | 2 | 1 | 5 |
| **Renal failure** | Post-COVID | <10 | 1.9 | 3 | 1 | 7 |
|  | 6 months prior | <10 | 1.9 | 1 | 0 | 4 |
|  | 12 months prior | 10 | 1.9 | 5 | 3 | 10 |
| **GORD** | Post-COVID | 36 | 1.8 | 19 | 14 | 27 |
|  | 6 months prior | 26 | 1.9 | 14 | 9 | 20 |
|  | 12 months prior | 32 | 1.9 | 17 | 12 | 24 |
| **Liver disease** | Post-COVID | 24 | 1.9 | 13 | 9 | 19 |
|  | 6 months prior | <10 | 1.9 | 5 | 2 | 9 |
|  | 12 months prior | 20 | 1.9 | 11 | 7 | 16 |
| **Diabetes** | Post-COVID | 35 | 1.8 | 19 | 14 | 27 |
|  | 6 months prior | <10 | 1.9 | 4 | 2 | 9 |
|  | 12 months prior | 31 | 1.9 | 17 | 12 | 24 |
| **Adrenal disease** | Post-COVID | <10 | 1.9 | 0 | n/a | n/a |
|  | 6 months prior | <10 | 1.9 | 0 | n/a | n/a |
|  | 12 months prior | <10 | 1.9 | 0 | n/a | n/a |
| **Arthritis** | Post-COVID | <10 | 1.9 | 2 | 1 | 5 |
|  | 6 months prior | <10 | 1.9 | 3 | 1 | 6 |
|  | 12 months prior | <10 | 1.9 | 5 | 2 | 9 |
| **Anxiety** | Post-COVID | 104 | 1.8 | 58 | 48 | 70 |
|  | 6 months prior | 61 | 1.8 | 33 | 26 | 43 |
|  | 12 months prior | 130 | 1.8 | 71 | 60 | 84 |
| **Depression** | Post-COVID | 67 | 1.8 | 37 | 29 | 47 |
|  | 6 months prior | 48 | 1.9 | 26 | 20 | 34 |
|  | 12 months prior | 103 | 1.9 | 55 | 46 | 67 |

### Table S12: Prescription event rates (per 100,000 person-weeks) in men 50 years old or younger with community COVID-19.

| **Outcome** | **Time point** | **Number of events** | **Time (person-weeks (PW))** | **Rate (per 100,000 PW)** | **Lower CI** | **Upper CI** |
| --- | --- | --- | --- | --- | --- | --- |
| **Diuretics** | Post-COVID | <10 | 1.9 | 3 | 1 | 6 |
|  | 6 months prior | <10 | 1.9 | 2 | 1 | 5 |
|  | 12 months prior | <10 | 1.9 | 2 | 1 | 6 |
| **Bronchodilators** | Post-COVID | 63 | 1.7 | 36 | 28 | 47 |
|  | 6 months prior | 68 | 1.8 | 38 | 30 | 49 |
|  | 12 months prior | 71 | 1.8 | 40 | 31 | 50 |
| **ICS** | Post-COVID | 47 | 1.8 | 27 | 20 | 36 |
|  | 6 months prior | 40 | 1.8 | 22 | 16 | 30 |
|  | 12 months prior | 46 | 1.8 | 25 | 19 | 34 |
| **Paracetamol** | Post-COVID | <10 | 1.9 | 1 | 0 | 4 |
|  | 6 months prior | <10 | 1.9 | 3 | 1 | 7 |
|  | 12 months prior | <10 | 1.9 | 4 | 2 | 8 |
| **NSAID** | Post-COVID | 61 | 1.8 | 35 | 27 | 44 |
|  | 6 months prior | 97 | 1.8 | 54 | 44 | 65 |
|  | 12 months prior | 101 | 1.8 | 56 | 46 | 68 |
| **Opioid medication** | Post-COVID | 56 | 1.8 | 31 | 24 | 41 |
|  | 6 months prior | 64 | 1.8 | 35 | 27 | 45 |
|  | 12 months prior | 84 | 1.8 | 46 | 37 | 57 |
| **Neuropathic pain medication** | Post-COVID | 45 | 1.8 | 25 | 18 | 33 |
|  | 6 months prior | 29 | 1.9 | 15 | 11 | 22 |
|  | 12 months prior | 36 | 1.9 | 19 | 14 | 27 |

### Table S13: Symptom event rates (per 100,000 person-weeks) in women over 50 years old with community COVID-19.

| **Outcome** | **Time point** | **Number of events** | **Time (person-weeks (PW))** | **Rate (per 100,000 PW)** | **Lower CI** | **Upper CI** |
| --- | --- | --- | --- | --- | --- | --- |
| **General pain** | Post-COVID | 14 | 0.9 | 16 | 10 | 27 |
|  | 6 months prior | <10 | 0.9 | 8 | 4 | 16 |
|  | 12 months prior | 17 | 0.9 | 19 | 12 | 31 |
| **Chest pain** | Post-COVID | 59 | 0.9 | 68 | 53 | 88 |
|  | 6 months prior | 33 | 0.9 | 37 | 26 | 52 |
|  | 12 months prior | 51 | 0.9 | 57 | 44 | 75 |
| **Abdominal pain** | Post-COVID | 72 | 0.9 | 83 | 66 | 105 |
|  | 6 months prior | 46 | 0.9 | 52 | 39 | 69 |
|  | 12 months prior | 50 | 0.9 | 56 | 43 | 75 |
| **Headache** | Post-COVID | 36 | 0.9 | 42 | 30 | 58 |
|  | 6 months prior | 29 | 0.9 | 33 | 23 | 47 |
|  | 12 months prior | 39 | 0.9 | 44 | 32 | 60 |
| **Joint pain** | Post-COVID | 254 | 0.8 | 306 | 270 | 345 |
|  | 6 months prior | 213 | 0.9 | 245 | 214 | 280 |
|  | 12 months prior | 280 | 0.9 | 326 | 290 | 367 |
| **Muscle pain** | Post-COVID | 14 | 0.9 | 16 | 10 | 27 |
|  | 6 months prior | <10 | 0.9 | 10 | 5 | 19 |
|  | 12 months prior | 13 | 0.9 | 15 | 8 | 25 |
| **Neuropathic pain** | Post-COVID | <10 | 0.9 | 0 | n/a | n/a |
|  | 6 months prior | <10 | 0.9 | 2 | 1 | 9 |
|  | 12 months prior | <10 | 0.9 | 6 | 2 | 13 |
| **Fatigue** | Post-COVID | 62 | 0.9 | 72 | 56 | 92 |
|  | 6 months prior | 25 | 0.9 | 28 | 19 | 42 |
|  | 12 months prior | 28 | 0.9 | 31 | 22 | 46 |
| **Fever** | Post-COVID | <10 | 0.9 | 9 | 5 | 19 |
|  | 6 months prior | 16 | 0.9 | 18 | 11 | 29 |
|  | 12 months prior | <10 | 0.9 | 10 | 5 | 19 |
| **Breathlessness** | Post-COVID | 133 | 0.9 | 156 | 132 | 185 |
|  | 6 months prior | 74 | 0.9 | 84 | 67 | 105 |
|  | 12 months prior | 81 | 0.9 | 92 | 74 | 114 |
| **Cough** | Post-COVID | 85 | 0.8 | 100 | 81 | 124 |
|  | 6 months prior | 77 | 0.9 | 88 | 70 | 110 |
|  | 12 months prior | 207 | 0.9 | 236 | 206 | 270 |
| **Chest tightness** | Post-COVID | <10 | 0.9 | 2 | 1 | 9 |
|  | 6 months prior | <10 | 0.9 | 1 | 0 | 8 |
|  | 12 months prior | <10 | 0.9 | 0 | n/a | n/a |
| **Palpitations** | Post-COVID | 28 | 0.9 | 32 | 22 | 47 |
|  | 6 months prior | 13 | 0.9 | 15 | 8 | 25 |
|  | 12 months prior | 22 | 0.9 | 25 | 16 | 37 |
| **Diarrhoea** | Post-COVID | 20 | 0.9 | 23 | 15 | 36 |
|  | 6 months prior | 14 | 0.9 | 16 | 9 | 26 |
|  | 12 months prior | 18 | 0.9 | 20 | 13 | 32 |
| **Nausea** | Post-COVID | <10 | 0.9 | 10 | 5 | 20 |
|  | 6 months prior | <10 | 0.9 | 8 | 4 | 16 |
|  | 12 months prior | <10 | 0.9 | 8 | 4 | 16 |
| **Anorexia** | Post-COVID | <10 | 0.9 | 8 | 4 | 17 |
|  | 6 months prior | <10 | 0.9 | 8 | 4 | 16 |
|  | 12 months prior | <10 | 0.9 | 7 | 3 | 15 |
| **Cognitive impairment** | Post-COVID | 11 | 0.9 | 13 | 7 | 23 |
|  | 6 months prior | 11 | 0.9 | 12 | 7 | 22 |
|  | 12 months prior | 19 | 0.9 | 21 | 14 | 33 |
| **Delirium** | Post-COVID | <10 | 0.9 | 10 | 5 | 20 |
|  | 6 months prior | 13 | 0.9 | 15 | 8 | 25 |
|  | 12 months prior | <10 | 0.9 | 6 | 2 | 13 |
| **Insomnia** | Post-COVID | 18 | 0.9 | 21 | 13 | 33 |
|  | 6 months prior | 12 | 0.9 | 13 | 8 | 24 |
|  | 12 months prior | 15 | 0.9 | 17 | 10 | 28 |
| **Dizziness** | Post-COVID | 35 | 0.9 | 41 | 29 | 56 |
|  | 6 months prior | 40 | 0.9 | 45 | 33 | 61 |
|  | 12 months prior | 36 | 0.9 | 41 | 29 | 56 |
| **Paranesthesia** | Post-COVID | <10 | 0.9 | 10 | 5 | 20 |
|  | 6 months prior | 13 | 0.9 | 15 | 8 | 25 |
|  | 12 months prior | 14 | 0.9 | 16 | 9 | 26 |
| **Earache** | Post-COVID | 25 | 0.9 | 29 | 19 | 43 |
|  | 6 months prior | 15 | 0.9 | 17 | 10 | 28 |
|  | 12 months prior | 23 | 0.9 | 26 | 17 | 39 |
| **Sore throat** | Post-COVID | <10 | 0.9 | 9 | 5 | 19 |
|  | 6 months prior | 22 | 0.9 | 25 | 16 | 37 |
|  | 12 months prior | 28 | 0.9 | 31 | 22 | 45 |
| **Smell/taste loss** | Post-COVID | <10 | 0.9 | 5 | 2 | 12 |
|  | 6 months prior | <10 | 0.9 | 7 | 3 | 15 |
| **Tinnitus** | Post-COVID | <10 | 0.9 | 10 | 5 | 20 |
|  | 6 months prior | <10 | 0.9 | 1 | 0 | 8 |
|  | 12 months prior | <10 | 0.9 | 7 | 3 | 15 |
| **Skin rash** | Post-COVID | 31 | 0.9 | 36 | 25 | 51 |
|  | 6 months prior | 39 | 0.9 | 44 | 32 | 60 |
|  | 12 months prior | 41 | 0.9 | 46 | 34 | 63 |

### Table S14: Disease outcome events (per 100,000 person-weeks) in women over 50 years old with community COVID-19.

| **Outcome** | **Time point** | **Number of events** | **Time (person-weeks (PW))** | **Rate (per 100,000 PW)** | **Lower CI** | **Upper CI** |
| --- | --- | --- | --- | --- | --- | --- |
| **Asthma** | Post-COVID | 60 | 0.8 | 76 | 59 | 98 |
|  | 6 months prior | 54 | 0.8 | 67 | 51 | 88 |
|  | 12 months prior | 69 | 0.8 | 85 | 67 | 108 |
| **Lung fibrosis** | Post-COVID | <10 | 0.9 | 5 | 2 | 12 |
|  | 6 months prior | <10 | 0.9 | 1 | 0 | 8 |
|  | 12 months prior | <10 | 0.9 | 1 | 0 | 8 |
| **Heart failure** | Post-COVID | <10 | 0.9 | 2 | 1 | 9 |
|  | 6 months prior | <10 | 0.9 | 9 | 5 | 18 |
|  | 12 months prior | <10 | 0.9 | 6 | 2 | 14 |
| **IHD** | Post-COVID | 13 | 0.9 | 15 | 9 | 26 |
|  | 6 months prior | 10 | 0.9 | 11 | 6 | 21 |
|  | 12 months prior | 14 | 0.9 | 16 | 9 | 27 |
| **Hypertension** | Post-COVID | 94 | 0.8 | 119 | 97 | 145 |
|  | 6 months prior | 54 | 0.8 | 67 | 51 | 88 |
|  | 12 months prior | 96 | 0.8 | 120 | 98 | 146 |
| **Stroke** | Post-COVID | 11 | 0.9 | 13 | 7 | 23 |
|  | 6 months prior | <10 | 0.9 | 8 | 4 | 17 |
|  | 12 months prior | 14 | 0.9 | 16 | 9 | 27 |
| **PAD** | Post-COVID | <10 | 0.9 | 0 | n/a | n/a |
|  | 6 months prior | <10 | 0.9 | 0 | n/a | n/a |
|  | 12 months prior | <10 | 0.9 | 0 | n/a | n/a |
| **Anemia** | Post-COVID | 22 | 0.9 | 26 | 17 | 39 |
|  | 6 months prior | 20 | 0.9 | 23 | 15 | 35 |
|  | 12 months prior | 24 | 0.9 | 27 | 18 | 41 |
| **VTE** | Post-COVID | 19 | 0.9 | 22 | 14 | 35 |
|  | 6 months prior | 14 | 0.9 | 16 | 9 | 27 |
|  | 12 months prior | 12 | 0.9 | 14 | 8 | 24 |
| **Renal failure** | Post-COVID | 27 | 0.9 | 32 | 22 | 46 |
|  | 6 months prior | 15 | 0.9 | 17 | 10 | 28 |
|  | 12 months prior | 21 | 0.9 | 24 | 16 | 37 |
| **GORD** | Post-COVID | 29 | 0.9 | 34 | 24 | 49 |
|  | 6 months prior | 24 | 0.9 | 27 | 18 | 41 |
|  | 12 months prior | 30 | 0.9 | 34 | 24 | 49 |
| **Liver disease** | Post-COVID | 17 | 0.9 | 20 | 12 | 32 |
|  | 6 months prior | 15 | 0.9 | 17 | 10 | 28 |
|  | 12 months prior | 18 | 0.9 | 20 | 13 | 32 |
| **Diabetes** | Post-COVID | 58 | 0.8 | 74 | 57 | 96 |
|  | 6 months prior | 23 | 0.8 | 29 | 19 | 43 |
|  | 12 months prior | 63 | 0.8 | 79 | 61 | 101 |
| **Adrenal disease** | Post-COVID | <10 | 0.9 | 1 | 0 | 8 |
|  | 6 months prior | <10 | 0.9 | 0 | n/a | n/a |
|  | 12 months prior | <10 | 0.9 | 0 | n/a | n/a |
| **Arthritis** | Post-COVID | 42 | 0.8 | 51 | 38 | 69 |
|  | 6 months prior | 25 | 0.8 | 30 | 20 | 44 |
|  | 12 months prior | 76 | 0.8 | 92 | 73 | 115 |
| **Anxiety** | Post-COVID | 71 | 0.8 | 86 | 68 | 109 |
|  | 6 months prior | 60 | 0.8 | 71 | 55 | 91 |
|  | 12 months prior | 76 | 0.8 | 90 | 72 | 112 |
| **Depression** | Post-COVID | 63 | 0.8 | 76 | 59 | 97 |
|  | 6 months prior | 35 | 0.9 | 41 | 29 | 57 |
|  | 12 months prior | 49 | 0.9 | 57 | 43 | 76 |

### Table S15: Prescription event rates (per 100,000 person-weeks) in women over 50 years old with community COVID-19.

| **Outcome** | **Time point** | **Number of events** | **Time (person-weeks (PW))** | **Rate (per 100,000 PW)** | **Lower CI** | **Upper CI** |
| --- | --- | --- | --- | --- | --- | --- |
| **Diuretics** | Post-COVID | <10 | 0.8 | 9 | 4 | 18 |
|  | 6 months prior | <10 | 0.8 | 7 | 3 | 16 |
|  | 12 months prior | 13 | 0.8 | 16 | 9 | 27 |
| **Bronchodilators** | Post-COVID | 62 | 0.7 | 85 | 67 | 110 |
|  | 6 months prior | 45 | 0.8 | 60 | 45 | 80 |
|  | 12 months prior | 69 | 0.8 | 91 | 72 | 115 |
| **ICS** | Post-COVID | 31 | 0.8 | 41 | 29 | 59 |
|  | 6 months prior | 33 | 0.8 | 42 | 30 | 60 |
|  | 12 months prior | 43 | 0.8 | 55 | 40 | 74 |
| **Paracetamol** | Post-COVID | 46 | 0.8 | 60 | 45 | 81 |
|  | 6 months prior | 51 | 0.8 | 65 | 50 | 86 |
|  | 12 months prior | 40 | 0.8 | 51 | 37 | 70 |
| **NSAID** | Post-COVID | 101 | 0.7 | 142 | 117 | 172 |
|  | 6 months prior | 92 | 0.7 | 127 | 104 | 156 |
|  | 12 months prior | 93 | 0.7 | 129 | 105 | 158 |
| **Opioid medication** | Post-COVID | 122 | 0.7 | 182 | 153 | 218 |
|  | 6 months prior | 105 | 0.7 | 153 | 127 | 186 |
|  | 12 months prior | 105 | 0.7 | 155 | 128 | 187 |
| **Neuropathic pain medication** | Post-COVID | 56 | 0.8 | 75 | 58 | 98 |
|  | 6 months prior | 55 | 0.8 | 72 | 55 | 94 |
|  | 12 months prior | 71 | 0.8 | 93 | 74 | 117 |

### Table S16: Symptom event rates (per 100,000 person-weeks) in women 50 years old or younger with community COVID-19.

| **Outcome** | **Time point** | **Number of events** | **Time (person-weeks (PW))** | **Rate (per 100,000 PW)** | **Lower CI** | **Upper CI** |
| --- | --- | --- | --- | --- | --- | --- |
| **General pain** | Post-COVID | 19 | 0.7 | 8 | 5 | 13 |
|  | 6 months prior | 11 | 0.8 | 5 | 3 | 9 |
|  | 12 months prior | 15 | 0.8 | 6 | 4 | 11 |
| **Chest pain** | Post-COVID | 108 | 2.3 | 48 | 40 | 58 |
|  | 6 months prior | 56 | 2.3 | 24 | 19 | 31 |
|  | 12 months prior | 62 | 2.3 | 27 | 21 | 34 |
| **Abdominal pain** | Post-COVID | 206 | 2.3 | 92 | 80 | 105 |
|  | 6 months prior | 152 | 2.3 | 66 | 56 | 77 |
|  | 12 months prior | 218 | 2.3 | 95 | 83 | 108 |
| **Headache** | Post-COVID | 182 | 2.2 | 81 | 70 | 94 |
|  | 6 months prior | 127 | 2.3 | 55 | 46 | 65 |
|  | 12 months prior | 172 | 2.3 | 75 | 64 | 87 |
| **Joint pain** | Post-COVID | 361 | 2.2 | 163 | 147 | 181 |
|  | 6 months prior | 316 | 2.3 | 138 | 123 | 154 |
|  | 12 months prior | 419 | 2.3 | 184 | 167 | 202 |
| **Muscle pain** | Post-COVID | 15 | 2.2 | 7 | 4 | 11 |
|  | 6 months prior | <10 | 2.3 | 2 | 1 | 5 |
|  | 12 months prior | <10 | 2.3 | 3 | 1 | 6 |
| **Neuropathic pain** | Post-COVID | <10 | 2.3 | 2 | 1 | 5 |
|  | 6 months prior | <10 | 2.3 | 1 | 0 | 4 |
|  | 12 months prior | <10 | 2.3 | 1 | 0 | 4 |
| **Fatigue** | Post-COVID | 124 | 2.3 | 55 | 46 | 66 |
|  | 6 months prior | 64 | 2.3 | 28 | 22 | 35 |
|  | 12 months prior | 124 | 2.3 | 54 | 45 | 64 |
| **Fever** | Post-COVID | 10 | 2.2 | 4 | 2 | 8 |
|  | 6 months prior | 28 | 2.3 | 12 | 8 | 17 |
|  | 12 months prior | 23 | 2.3 | 10 | 7 | 15 |
| **Breathlessness** | Post-COVID | 157 | 2.3 | 70 | 60 | 82 |
|  | 6 months prior | 68 | 2.3 | 29 | 23 | 37 |
|  | 12 months prior | 41 | 2.3 | 18 | 13 | 24 |
| **Cough** | Post-COVID | 106 | 2.2 | 47 | 39 | 57 |
|  | 6 months prior | 90 | 2.3 | 39 | 32 | 48 |
|  | 12 months prior | 232 | 2.3 | 100 | 88 | 114 |
| **Chest tightness** | Post-COVID | 14 | 2.2 | 6 | 4 | 10 |
|  | 6 months prior | 10 | 2.3 | 4 | 2 | 8 |
|  | 12 months prior | 10 | 2.3 | 4 | 2 | 8 |
| **Palpitations** | Post-COVID | 42 | 2.3 | 19 | 14 | 25 |
|  | 6 months prior | 26 | 2.3 | 11 | 8 | 16 |
|  | 12 months prior | 27 | 2.3 | 12 | 8 | 17 |
| **Diarrhea** | Post-COVID | 19 | 2.3 | 8 | 5 | 13 |
|  | 6 months prior | 26 | 2.3 | 11 | 8 | 16 |
|  | 12 months prior | 30 | 2.3 | 13 | 9 | 18 |
| **Nausea** | Post-COVID | 22 | 2.3 | 10 | 6 | 15 |
|  | 6 months prior | 18 | 2.3 | 8 | 5 | 12 |
|  | 12 months prior | 16 | 2.3 | 7 | 4 | 11 |
| **Anorexia** | Post-COVID | <10 | 2.3 | 4 | 2 | 7 |
|  | 6 months prior | <10 | 2.3 | 2 | 1 | 5 |
|  | 12 months prior | <10 | 2.3 | 3 | 2 | 7 |
| **Cognitive impairment** | Post-COVID | <10 | 2.3 | 3 | 1 | 6 |
|  | 6 months prior | <10 | 2.3 | 0 | 0 | 3 |
|  | 12 months prior | <10 | 2.3 | 4 | 2 | 7 |
| **Delirium** | Post-COVID | <10 | 2.3 | 1 | 0 | 4 |
|  | 6 months prior | <10 | 2.3 | 1 | 0 | 4 |
|  | 12 months prior | <10 | 2.3 | 1 | 0 | 3 |
| **Insomnia** | Post-COVID | 33 | 2.3 | 15 | 10 | 21 |
|  | 6 months prior | 21 | 2.3 | 9 | 6 | 14 |
|  | 12 months prior | 25 | 2.3 | 11 | 7 | 16 |
| **Dizziness** | Post-COVID | 51 | 2.3 | 23 | 17 | 30 |
|  | 6 months prior | 46 | 2.3 | 20 | 15 | 26 |
|  | 12 months prior | 77 | 2.3 | 33 | 27 | 41 |
| **Paranesthesia** | Post-COVID | 22 | 2.3 | 10 | 6 | 15 |
|  | 6 months prior | 17 | 2.3 | 7 | 5 | 12 |
|  | 12 months prior | 21 | 2.3 | 9 | 6 | 14 |
| **Earache** | Post-COVID | 41 | 2.3 | 18 | 13 | 25 |
|  | 6 months prior | 28 | 2.3 | 12 | 8 | 17 |
|  | 12 months prior | 39 | 2.3 | 17 | 12 | 23 |
| **Sore throat** | Post-COVID | 65 | 2.3 | 29 | 23 | 37 |
|  | 6 months prior | 62 | 2.3 | 27 | 21 | 34 |
|  | 12 months prior | 171 | 2.3 | 74 | 64 | 86 |
| **Smell/taste loss** | Post-COVID | 17 | 2.2 | 8 | 5 | 12 |
|  | 6 months prior | 16 | 2.3 | 7 | 4 | 11 |
| **Tinnitus** | Post-COVID | 12 | 2.3 | 5 | 3 | 9 |
|  | 6 months prior | <10 | 2.3 | 2 | 1 | 5 |
|  | 12 months prior | <10 | 2.3 | 4 | 2 | 7 |
| **Skin rash** | Post-COVID | 119 | 2.3 | 53 | 44 | 63 |
|  | 6 months prior | 88 | 2.3 | 38 | 31 | 47 |
|  | 12 months prior | 168 | 2.3 | 73 | 62 | 84 |

### Table S17: Disease outcome events (per 100,000 person-weeks) in women 50 years old or younger with community COVID-19.

| **Outcome** | **Time point** | **Number of events** | **Time (person-weeks (PW))** | **Rate (per 100,000 PW)** | **Lower CI** | **Upper CI** |
| --- | --- | --- | --- | --- | --- | --- |
| **Asthma** | Post-COVID | 126 | 2.1 | 59 | 50 | 71 |
|  | 6 months prior | 143 | 2.2 | 65 | 55 | 77 |
|  | 12 months prior | 135 | 2.2 | 61 | 52 | 73 |
| **Lung fibrosis** | Post-COVID | <10 | 2.3 | 0 | 0 | 3 |
|  | 6 months prior | <10 | 2.3 | 0 | n/a | n/a |
|  | 12 months prior | <10 | 2.3 | 0 | 0 | 3 |
| **Heart failure** | Post-COVID | <10 | 2.3 | 0 | 0 | 3 |
|  | 6 months prior | <10 | 2.3 | 0 | n/a | n/a |
|  | 12 months prior | <10 | 2.3 | 1 | 0 | 3 |
| **IHD** | Post-COVID | <10 | 2.3 | 2 | 1 | 5 |
|  | 6 months prior | <10 | 2.3 | 0 | n/a | n/a |
|  | 12 months prior | <10 | 2.3 | 2 | 1 | 5 |
| **Hypertension** | Post-COVID | 44 | 2.2 | 20 | 15 | 26 |
|  | 6 months prior | 19 | 2.3 | 8 | 5 | 13 |
|  | 12 months prior | 35 | 2.3 | 15 | 11 | 21 |
| **Stroke** | Post-COVID | <10 | 2.3 | 1 | 0 | 4 |
|  | 6 months prior | <10 | 2.3 | 0 | 0 | 3 |
|  | 12 months prior | <10 | 2.3 | 2 | 1 | 5 |
| **PAD** | Post-COVID | <10 | 2.3 | 0 | n/a | n/a |
|  | 6 months prior | <10 | 2.3 | 0 | 0 | 3 |
|  | 12 months prior | <10 | 2.3 | 0 | 0 | 3 |
| **Anaemia** | Post-COVID | 43 | 2.2 | 19 | 14 | 26 |
|  | 6 months prior | 19 | 2.3 | 8 | 5 | 13 |
|  | 12 months prior | 62 | 2.3 | 27 | 21 | 35 |
| **VTE** | Post-COVID | 15 | 2.3 | 7 | 4 | 11 |
|  | 6 months prior | <10 | 2.3 | 1 | 0 | 4 |
|  | 12 months prior | <10 | 2.3 | 3 | 1 | 6 |
| **Renal failure** | Post-COVID | 12 | 2.3 | 5 | 3 | 9 |
|  | 6 months prior | <10 | 2.3 | 3 | 1 | 6 |
|  | 12 months prior | 11 | 2.3 | 5 | 3 | 9 |
| **GORD** | Post-COVID | 41 | 2.2 | 18 | 14 | 25 |
|  | 6 months prior | 47 | 2.3 | 20 | 15 | 27 |
|  | 12 months prior | 50 | 2.3 | 22 | 16 | 29 |
| **Liver disease** | Post-COVID | 17 | 2.3 | 8 | 5 | 12 |
|  | 6 months prior | <10 | 2.3 | 2 | 1 | 5 |
|  | 12 months prior | 16 | 2.3 | 7 | 4 | 11 |
| **Diabetes** | Post-COVID | 42 | 2.2 | 19 | 14 | 26 |
|  | 6 months prior | 25 | 2.3 | 11 | 7 | 16 |
|  | 12 months prior | 47 | 2.3 | 21 | 15 | 27 |
| **Adrenal disease** | Post-COVID | <10 | 2.3 | 0 | n/a | n/a |
|  | 6 months prior | <10 | 2.3 | 0 | n/a | n/a |
|  | 12 months prior | <10 | 2.3 | 0 | n/a | n/a |
| **Arthritis** | Post-COVID | 17 | 2.2 | 8 | 5 | 12 |
|  | 6 months prior | <10 | 2.3 | 3 | 2 | 7 |
|  | 12 months prior | 23 | 2.3 | 10 | 7 | 15 |
| **Anxiety** | Post-COVID | 303 | 2.1 | 147 | 132 | 165 |
|  | 6 months prior | 200 | 2.1 | 94 | 82 | 109 |
|  | 12 months prior | 270 | 2.1 | 127 | 113 | 143 |
| **Depression** | Post-COVID | 189 | 2.1 | 89 | 77 | 103 |
|  | 6 months prior | 129 | 2.2 | 59 | 50 | 70 |
|  | 12 months prior | 183 | 2.2 | 83 | 72 | 96 |

### Table S18: Prescription event rates (per 100,000 person-weeks) in women 50 years old or younger with community COVID-19.

| **Outcome** | **Time point** | **Number of events** | **Time (person-weeks (PW))** | **Rate (per 100,000 PW)** | **Lower CI** | **Upper CI** |
| --- | --- | --- | --- | --- | --- | --- |
| **Diuretics** | Post-COVID | <10 | 2.3 | 1 | 0 | 4 |
|  | 6 months prior | <10 | 2.3 | 1 | 0 | 3 |
|  | 12 months prior | <10 | 2.3 | 1 | 0 | 4 |
| **Bronchodilators** | Post-COVID | 142 | 2.0 | 69 | 59 | 82 |
|  | 6 months prior | 95 | 2.1 | 45 | 37 | 55 |
|  | 12 months prior | 133 | 2.1 | 62 | 52 | 74 |
| **ICS** | Post-COVID | 72 | 2.1 | 34 | 27 | 43 |
|  | 6 months prior | 59 | 2.2 | 27 | 21 | 35 |
|  | 12 months prior | 68 | 2.2 | 31 | 24 | 39 |
| **Paracetamol** | Post-COVID | 18 | 2.2 | 8 | 5 | 13 |
|  | 6 months prior | 18 | 2.3 | 8 | 5 | 12 |
|  | 12 months prior | 18 | 2.3 | 8 | 5 | 12 |
| **NSAID** | Post-COVID | 206 | 2.1 | 100 | 87 | 115 |
|  | 6 months prior | 190 | 2.1 | 90 | 78 | 104 |
|  | 12 months prior | 198 | 2.1 | 94 | 81 | 108 |
| **Opioid medication** | Post-COVID | 160 | 2.1 | 78 | 67 | 91 |
|  | 6 months prior | 152 | 2.1 | 72 | 61 | 84 |
|  | 12 months prior | 166 | 2.1 | 79 | 67 | 91 |
| **Neuropathic pain medication** | Post-COVID | 110 | 2.1 | 51 | 42 | 62 |
|  | 6 months prior | 76 | 2.2 | 34 | 27 | 43 |
|  | 12 months prior | 85 | 2.2 | 38 | 31 | 47 |

### Table S19: Symptom event rates (per 100,000 person-weeks) for events identified from 2 weeks after COVID-19 diagnosis.

| **Outcome** | **Time point** | **COVID exposure group** | **Number of events** | **Time (person-weeks (PW))** | **Rate (per 100,000 PW)** | **Lower CI** | **Upper CI** |
| --- | --- | --- | --- | --- | --- | --- | --- |
| **General pain** | Post-COVID | Community | 55 | 5.7 | 10 | 7 | 13 |
|  |  | Hospital | <10 | 0.2 | 18 | 6 | 55 |
|  | 6 months prior | Community | 37 | 5.9 | 6 | 5 | 9 |
|  |  | Hospital | <10 | 0.2 | 5 | 1 | 39 |
|  | 12 months prior | Community | 55 | 5.9 | 9 | 7 | 12 |
|  |  | Hospital | <10 | 0.2 | 22 | 8 | 58 |
| **Chest pain** | Post-COVID | Community | 349 | 5.7 | 61 | 55 | 68 |
|  |  | Hospital | 29 | 0.2 | 175 | 122 | 253 |
|  | 6 months prior | Community | 180 | 5.9 | 31 | 27 | 36 |
|  |  | Hospital | 13 | 0.2 | 72 | 42 | 124 |
|  | 12 months prior | Community | 247 | 5.9 | 42 | 37 | 48 |
|  |  | Hospital | 16 | 0.2 | 88 | 54 | 144 |
| **Abdominal pain** | Post-COVID | Community | 457 | 5.7 | 81 | 73 | 88 |
|  |  | Hospital | 21 | 0.2 | 127 | 83 | 195 |
|  | 6 months prior | Community | 324 | 5.8 | 55 | 50 | 62 |
|  |  | Hospital | 25 | 0.2 | 140 | 95 | 207 |
|  | 12 months prior | Community | 451 | 5.8 | 77 | 71 | 85 |
|  |  | Hospital | 25 | 0.2 | 138 | 93 | 204 |
| **Headache** | Post-COVID | Community | 367 | 5.7 | 65 | 59 | 72 |
|  |  | Hospital | 16 | 0.2 | 97 | 59 | 158 |
|  | 6 months prior | Community | 262 | 5.8 | 45 | 40 | 51 |
|  |  | Hospital | 13 | 0.2 | 72 | 42 | 125 |
|  | 12 months prior | Community | 361 | 5.8 | 62 | 56 | 69 |
|  |  | Hospital | 15 | 0.2 | 82 | 50 | 136 |
| **Joint pain** | Post-COVID | Community | 1108 | 5.6 | 199 | 188 | 211 |
|  |  | Hospital | 58 | 0.2 | 366 | 283 | 474 |
|  | 6 months prior | Community | 992 | 5.7 | 173 | 162 | 184 |
|  |  | Hospital | 57 | 0.2 | 332 | 256 | 431 |
|  | 12 months prior | Community | 1453 | 5.7 | 255 | 242 | 268 |
|  |  | Hospital | 80 | 0.2 | 459 | 369 | 571 |
| **Muscle pain** | Post-COVID | Community | 62 | 5.7 | 11 | 8 | 14 |
|  |  | Hospital | <10 | 0.2 | 35 | 16 | 79 |
|  | 6 months prior | Community | 32 | 5.9 | 5 | 4 | 8 |
|  |  | Hospital | <10 | 0.2 | 16 | 5 | 51 |
|  | 12 months prior | Community | 47 | 5.9 | 8 | 6 | 11 |
|  |  | Hospital | <10 | 0.2 | 5 | 1 | 39 |
| **Neuropathic pain** | Post-COVID | Community | 10 | 5.7 | 2 | 1 | 3 |
|  |  | Hospital | <10 | 0.2 | 6 | 1 | 42 |
|  | 6 months prior | Community | <10 | 5.9 | 2 | 1 | 3 |
|  |  | Hospital | <10 | 0.2 | 11 | 3 | 44 |
|  | 12 months prior | Community | 13 | 5.9 | 2 | 1 | 4 |
|  |  | Hospital | <10 | 0.2 | 0 | n/a | n/a |
| **Fatigue** | Post-COVID | Community | 324 | 5.7 | 57 | 51 | 64 |
|  |  | Hospital | 26 | 0.2 | 156 | 106 | 229 |
|  | 6 months prior | Community | 132 | 5.9 | 23 | 19 | 27 |
|  |  | Hospital | <10 | 0.2 | 33 | 15 | 74 |
|  | 12 months prior | Community | 244 | 5.9 | 42 | 37 | 47 |
|  |  | Hospital | 10 | 0.2 | 55 | 29 | 102 |
| **Fever** | Post-COVID | Community | 56 | 5.7 | 10 | 8 | 13 |
|  |  | Hospital | <10 | 0.2 | 42 | 20 | 89 |
|  | 6 months prior | Community | 77 | 5.8 | 13 | 11 | 16 |
|  |  | Hospital | <10 | 0.2 | 17 | 5 | 52 |
|  | 12 months prior | Community | 51 | 5.9 | 9 | 7 | 11 |
|  |  | Hospital | <10 | 0.2 | 11 | 3 | 43 |
| **Breathlessness** | Post-COVID | Community | 479 | 5.7 | 85 | 77 | 93 |
|  |  | Hospital | 84 | 0.2 | 536 | 432 | 663 |
|  | 6 months prior | Community | 296 | 5.8 | 51 | 45 | 57 |
|  |  | Hospital | 42 | 0.2 | 243 | 180 | 329 |
|  | 12 months prior | Community | 261 | 5.9 | 45 | 39 | 50 |
|  |  | Hospital | 40 | 0.2 | 223 | 163 | 304 |
| **Cough** | Post-COVID | Community | 418 | 5.6 | 74 | 67 | 82 |
|  |  | Hospital | 32 | 0.2 | 202 | 143 | 285 |
|  | 6 months prior | Community | 358 | 5.8 | 62 | 56 | 69 |
|  |  | Hospital | 23 | 0.2 | 134 | 89 | 201 |
|  | 12 months prior | Community | 800 | 5.8 | 138 | 128 | 148 |
|  |  | Hospital | 58 | 0.2 | 324 | 251 | 420 |
| **Chest tightness** | Post-COVID | Community | 32 | 5.7 | 6 | 4 | 8 |
|  |  | Hospital | <10 | 0.2 | 0 | n/a | n/a |
|  | 6 months prior | Community | 23 | 5.9 | 4 | 3 | 6 |
|  |  | Hospital | <10 | 0.2 | 5 | 1 | 39 |
|  | 12 months prior | Community | 20 | 5.9 | 3 | 2 | 5 |
|  |  | Hospital | <10 | 0.2 | 0 | n/a | n/a |
| **Palpitations** | Post-COVID | Community | 122 | 5.7 | 21 | 18 | 26 |
|  |  | Hospital | <10 | 0.2 | 18 | 6 | 55 |
|  | 6 months prior | Community | 59 | 5.9 | 10 | 8 | 13 |
|  |  | Hospital | <10 | 0.2 | 38 | 18 | 80 |
|  | 12 months prior | Community | 83 | 5.9 | 14 | 11 | 18 |
|  |  | Hospital | <10 | 0.2 | 16 | 5 | 51 |
| **Diarrhea** | Post-COVID | Community | 83 | 5.7 | 15 | 12 | 18 |
|  |  | Hospital | 10 | 0.2 | 59 | 32 | 111 |
|  | 6 months prior | Community | 73 | 5.9 | 12 | 10 | 16 |
|  |  | Hospital | <10 | 0.2 | 38 | 18 | 81 |
|  | 12 months prior | Community | 97 | 5.9 | 17 | 14 | 20 |
|  |  | Hospital | 11 | 0.2 | 60 | 33 | 109 |
| **Nausea** | Post-COVID | Community | 41 | 5.7 | 7 | 5 | 10 |
|  |  | Hospital | <10 | 0.2 | 53 | 28 | 102 |
|  | 6 months prior | Community | 39 | 5.9 | 7 | 5 | 9 |
|  |  | Hospital | <10 | 0.2 | 22 | 8 | 58 |
|  | 12 months prior | Community | 34 | 5.9 | 6 | 4 | 8 |
|  |  | Hospital | <10 | 0.2 | 22 | 8 | 58 |
| **Anorexia** | Post-COVID | Community | 29 | 5.7 | 5 | 4 | 7 |
|  |  | Hospital | <10 | 0.2 | 24 | 9 | 63 |
|  | 6 months prior | Community | 22 | 5.9 | 4 | 2 | 6 |
|  |  | Hospital | <10 | 0.2 | 22 | 8 | 58 |
|  | 12 months prior | Community | 27 | 5.9 | 5 | 3 | 7 |
|  |  | Hospital | <10 | 0.2 | 5 | 1 | 39 |
| **Cognitive impairment** | Post-COVID | Community | 28 | 5.7 | 5 | 3 | 7 |
|  |  | Hospital | <10 | 0.2 | 18 | 6 | 55 |
|  | 6 months prior | Community | 22 | 5.9 | 4 | 2 | 6 |
|  |  | Hospital | <10 | 0.2 | 22 | 8 | 58 |
|  | 12 months prior | Community | 46 | 5.9 | 8 | 6 | 10 |
|  |  | Hospital | <10 | 0.2 | 38 | 18 | 80 |
| **Delirium** | Post-COVID | Community | 18 | 5.7 | 3 | 2 | 5 |
|  |  | Hospital | <10 | 0.2 | 30 | 12 | 71 |
|  | 6 months prior | Community | 20 | 5.9 | 3 | 2 | 5 |
|  |  | Hospital | <10 | 0.2 | 28 | 11 | 66 |
|  | 12 months prior | Community | 14 | 5.9 | 2 | 1 | 4 |
|  |  | Hospital | <10 | 0.2 | 33 | 15 | 73 |
| **Insomnia** | Post-COVID | Community | 92 | 5.7 | 16 | 13 | 20 |
|  |  | Hospital | <10 | 0.2 | 30 | 12 | 71 |
|  | 6 months prior | Community | 54 | 5.9 | 9 | 7 | 12 |
|  |  | Hospital | <10 | 0.2 | 49 | 26 | 95 |
|  | 12 months prior | Community | 69 | 5.9 | 12 | 9 | 15 |
|  |  | Hospital | <10 | 0.2 | 27 | 11 | 65 |
| **Dizziness** | Post-COVID | Community | 150 | 5.7 | 26 | 22 | 31 |
|  |  | Hospital | 12 | 0.2 | 71 | 41 | 126 |
|  | 6 months prior | Community | 144 | 5.9 | 25 | 21 | 29 |
|  |  | Hospital | 14 | 0.2 | 77 | 46 | 130 |
|  | 12 months prior | Community | 191 | 5.9 | 33 | 28 | 38 |
|  |  | Hospital | 14 | 0.2 | 77 | 45 | 129 |
| **Paresthesia** | Post-COVID | Community | 56 | 5.7 | 10 | 8 | 13 |
|  |  | Hospital | <10 | 0.2 | 18 | 6 | 55 |
|  | 6 months prior | Community | 53 | 5.9 | 9 | 7 | 12 |
|  |  | Hospital | <10 | 0.2 | 5 | 1 | 39 |
|  | 12 months prior | Community | 65 | 5.9 | 11 | 9 | 14 |
|  |  | Hospital | <10 | 0.2 | 27 | 11 | 65 |
| **Earache** | Post-COVID | Community | 104 | 5.7 | 18 | 15 | 22 |
|  |  | Hospital | <10 | 0.2 | 24 | 9 | 63 |
|  | 6 months prior | Community | 88 | 5.9 | 15 | 12 | 18 |
|  |  | Hospital | <10 | 0.2 | 49 | 26 | 94 |
|  | 12 months prior | Community | 100 | 5.9 | 17 | 14 | 21 |
|  |  | Hospital | <10 | 0.2 | 16 | 5 | 51 |
| **Sore throat** | Post-COVID | Community | 161 | 5.7 | 28 | 24 | 33 |
|  |  | Hospital | <10 | 0.2 | 18 | 6 | 55 |
|  | 6 months prior | Community | 170 | 5.8 | 29 | 25 | 34 |
|  |  | Hospital | 10 | 0.2 | 55 | 30 | 102 |
|  | 12 months prior | Community | 379 | 5.8 | 65 | 59 | 72 |
|  |  | Hospital | <10 | 0.2 | 22 | 8 | 58 |
| **Smell/taste loss** | Post-COVID | Community | 65 | 5.7 | 11 | 9 | 15 |
|  |  | Hospital | <10 | 0.2 | 35 | 16 | 79 |
|  | 6 months prior | Community | 36 | 5.9 | 6 | 4 | 9 |
|  |  | Hospital | <10 | 0.2 | 5 | 1 | 39 |
|  | 12 months prior* | Community | <10 | n/a | n/a | n/a | n/a |
|  |  | Hospital | <10 | n/a | n/a | n/a | n/a |
| **Tinnitus** | Post-COVID | Community | 42 | 5.7 | 7 | 5 | 10 |
|  |  | Hospital | <10 | 0.2 | 0 | n/a | n/a |
|  | 6 months prior | Community | 24 | 5.9 | 4 | 3 | 6 |
|  |  | Hospital | <10 | 0.2 | 11 | 3 | 44 |
|  | 12 months prior | Community | 36 | 5.9 | 6 | 4 | 8 |
|  |  | Hospital | <10 | 0.2 | 0 | n/a | n/a |
| **Skin rash** | Post-COVID | Community | 253 | 5.7 | 44 | 39 | 50 |
|  |  | Hospital | 14 | 0.2 | 83 | 49 | 140 |
|  | 6 months prior | Community | 232 | 5.9 | 40 | 35 | 45 |
|  |  | Hospital | 10 | 0.2 | 55 | 29 | 102 |
|  | 12 months prior | Community | 403 | 5.8 | 69 | 63 | 76 |
|  |  | Hospital | 17 | 0.2 | 93 | 58 | 150 |

*****No events for smell/taste loss using 12-month cohort.

### Table S20: Disease event rates (per 100,000 person-weeks) for events identified from 2 weeks after COVID-19 diagnosis.

| **Outcome** | **Time point** | **COVID exposure group** | **Number of events** | **Time (person-weeks (PW))** | **Rate (per 100,000 PW)** | **Lower CI** | **Upper CI** |
| --- | --- | --- | --- | --- | --- | --- | --- |
| **Asthma** | Post-COVID | Community | 322 | 5.3 | 60 | 54 | 67 |
|  |  | Hospital | 15 | 0.1 | 100 | 61 | 175 |
|  | 6 months prior | Community | 339 | 5.5 | 62 | 55 | 69 |
|  |  | Hospital | 14 | 0.2 | 87 | 51 | 146 |
|  | 12 months prior | Community | 358 | 5.5 | 65 | 58 | 72 |
|  |  | Hospital | 21 | 0.2 | 128 | 84 | 197 |
| **Lung fibrosis** | Post-COVID | Community | 16 | 5.7 | 3 | 2 | 5 |
|  |  | Hospital | <10 | 0.2 | 0 | n/a | n/a |
|  | 6 months prior | Community | <10 | 5.9 | 0 | 0 | 1 |
|  |  | Hospital | <10 | 0.2 | 5 | 1 | 39 |
|  | 12 months prior | Community | <10 | 5.9 | 1 | 0 | 2 |
|  |  | Hospital | <10 | 0.2 | 0 | n/a | n/a |
| **Heart failure** | Post-COVID | Community | 14 | 5.7 | 2 | 1 | 4 |
|  |  | Hospital | <10 | 0.2 | 54 | 29 | 116 |
|  | 6 months prior | Community | 16 | 5.9 | 3 | 2 | 4 |
|  |  | Hospital | <10 | 0.2 | 28 | 12 | 67 |
|  | 12 months prior | Community | 21 | 5.9 | 4 | 2 | 5 |
|  |  | Hospital | <10 | 0.2 | 28 | 12 | 67 |
| **IHD** | Post-COVID | Community | 48 | 5.7 | 8 | 6 | 11 |
|  |  | Hospital | 11 | 0.2 | 67 | 38 | 131 |
|  | 6 months prior | Community | 28 | 5.8 | 5 | 3 | 7 |
|  |  | Hospital | <10 | 0.2 | 22 | 8 | 60 |
|  | 12 months prior | Community | 59 | 5.8 | 10 | 8 | 13 |
|  |  | Hospital | <10 | 0.2 | 28 | 12 | 67 |
| **Hypertension** | Post-COVID | Community | 257 | 5.5 | 47 | 41 | 53 |
|  |  | Hospital | 36 | 0.1 | 244 | 178 | 344 |
|  | 6 months prior | Community | 138 | 5.6 | 24 | 21 | 29 |
|  |  | Hospital | 10 | 0.2 | 63 | 34 | 117 |
|  | 12 months prior | Community | 266 | 5.7 | 47 | 42 | 53 |
|  |  | Hospital | 34 | 0.2 | 211 | 151 | 296 |
| **Stroke** | Post-COVID | Community | 31 | 5.7 | 5 | 4 | 8 |
|  |  | Hospital | <10 | 0.2 | 37 | 17 | 96 |
|  | 6 months prior | Community | 20 | 5.9 | 3 | 2 | 5 |
|  |  | Hospital | <10 | 0.2 | 6 | 1 | 40 |
|  | 12 months prior | Community | 37 | 5.9 | 6 | 5 | 9 |
|  |  | Hospital | 12 | 0.2 | 67 | 38 | 118 |
| **PAD** | Post-COVID | Community | <10 | 5.7 | 0 | 0 | 4 |
|  |  | Hospital | <10 | 0.2 | 6 | n/a | n/a |
|  | 6 months prior | Community | <10 | 5.9 | 0 | 0 | 1 |
|  |  | Hospital | <10 | 0.2 | 5 | 1 | 39 |
|  | 12 months prior | Community | <10 | 5.9 | 1 | 1 | 3 |
|  |  | Hospital | <10 | 0.2 | 16 | 5 | 51 |
| **Anemia** | Post-COVID | Community | 83 | 5.7 | 15 | 12 | 18 |
|  |  | Hospital | 14 | 0.2 | 87 | 52 | 155 |
|  | 6 months prior | Community | 49 | 5.8 | 8 | 6 | 11 |
|  |  | Hospital | <10 | 0.2 | 46 | 23 | 91 |
|  | 12 months prior | Community | 108 | 5.8 | 19 | 15 | 22 |
|  |  | Hospital | 19 | 0.2 | 107 | 68 | 168 |
| **VTE** | Post-COVID | Community | 56 | 5.7 | 10 | 8 | 13 |
|  |  | Hospital | 30 | 0.2 | 185 | 130 | 271 |
|  | 6 months prior | Community | 37 | 5.9 | 6 | 5 | 9 |
|  |  | Hospital | <10 | 0.2 | 22 | 8 | 59 |
|  | 12 months prior | Community | 28 | 5.9 | 5 | 3 | 7 |
|  |  | Hospital | <10 | 0.2 | 11 | 3 | 44 |
| **Renal failure** | Post-COVID | Community | 62 | 5.7 | 11 | 9 | 14 |
|  |  | Hospital | 24 | 0.2 | 149 | 101 | 230 |
|  | 6 months prior | Community | 33 | 5.8 | 6 | 4 | 8 |
|  |  | Hospital | <10 | 0.2 | 23 | 9 | 60 |
|  | 12 months prior | Community | 63 | 5.8 | 11 | 8 | 14 |
|  |  | Hospital | <10 | 0.2 | 51 | 27 | 99 |
| **GORD** | Post-COVID | Community | 131 | 5.6 | 23 | 20 | 28 |
|  |  | Hospital | <10 | 0.2 | 18 | 6 | 88 |
|  | 6 months prior | Community | 120 | 5.8 | 21 | 17 | 25 |
|  |  | Hospital | <10 | 0.2 | 39 | 18 | 81 |
|  | 12 months prior | Community | 128 | 5.8 | 22 | 19 | 26 |
|  |  | Hospital | <10 | 0.2 | 22 | 8 | 59 |
| **Liver disease** | Post-COVID | Community | 77 | 5.7 | 14 | 11 | 17 |
|  |  | Hospital | <10 | 0.2 | 30 | 13 | 90 |
|  | 6 months prior | Community | 39 | 5.8 | 7 | 5 | 9 |
|  |  | Hospital | <10 | 0.2 | 17 | 5 | 52 |
|  | 12 months prior | Community | 69 | 5.8 | 12 | 9 | 15 |
|  |  | Hospital | <10 | 0.2 | 44 | 22 | 89 |
| **Diabetes** | Post-COVID | Community | 198 | 5.4 | 36 | 32 | 42 |
|  |  | Hospital | 42 | 0.1 | 303 | 225 | 416 |
|  | 6 months prior | Community | 82 | 5.6 | 15 | 12 | 18 |
|  |  | Hospital | <10 | 0.1 | 61 | 32 | 116 |
|  | 12 months prior | Community | 205 | 5.6 | 37 | 32 | 42 |
|  |  | Hospital | 19 | 0.1 | 128 | 81 | 200 |
| **Adrenal failure** | Post-COVID | Community | <10 | 5.7 | 0 | n/a | n/a |
|  |  | Hospital | <10 | 0.2 | 6 | n/a | n/a |
|  | 6 months prior | Community | <10 | 5.9 | 0 | n/a | n/a |
|  |  | Hospital | <10 | 0.2 | 0 | n/a | n/a |
|  | 12 months prior | Community | <10 | 5.9 | 0 | 0 | 1 |
|  |  | Hospital | <10 | 0.2 | 0 | n/a | n/a |
| **Arthritis** | Post-COVID | Community | 93 | 5.6 | 17 | 14 | 20 |
|  |  | Hospital | <10 | 0.2 | 51 | 26 | 113 |
|  | 6 months prior | Community | 59 | 5.8 | 10 | 8 | 13 |
|  |  | Hospital | <10 | 0.2 | 53 | 27 | 101 |
|  | 12 months prior | Community | 162 | 5.8 | 28 | 24 | 33 |
|  |  | Hospital | 13 | 0.2 | 76 | 44 | 132 |
| **Anxiety** | Post-COVID | Community | 502 | 5.4 | 93 | 86 | 102 |
|  |  | Hospital | 16 | 0.2 | 100 | 62 | 171 |
|  | 6 months prior | Community | 347 | 5.5 | 63 | 57 | 70 |
|  |  | Hospital | 13 | 0.2 | 74 | 43 | 128 |
|  | 12 months prior | Community | 495 | 5.5 | 90 | 82 | 98 |
|  |  | Hospital | 12 | 0.2 | 69 | 39 | 122 |
| **Depression** | Post-COVID | Community | 346 | 5.5 | 63 | 57 | 70 |
|  |  | Hospital | <10 | 0.2 | 55 | 29 | 117 |
|  | 6 months prior | Community | 223 | 5.6 | 40 | 35 | 45 |
|  |  | Hospital | <10 | 0.2 | 46 | 23 | 91 |
|  | 12 months prior | Community | 358 | 5.6 | 63 | 57 | 70 |
|  |  | Hospital | 10 | 0.2 | 57 | 31 | 106 |

### Table S21: Prescription event rates (per 100,000 person-weeks) for events identified from 2 weeks after COVID-19 diagnosis.

| **Outcome** | **Time point** | **COVID exposure group** | **Number of events** | **Time (person-weeks (PW))** | **Rate (per 100,000 PW)** | **Lower CI** | **Upper CI** |
| --- | --- | --- | --- | --- | --- | --- | --- |
| **Diuretics** | Post-COVID | Community | 23 | 5.6 | 4 | 3 | 6 |
|  |  | Hospital | <10 | 0.2 | 32 | 13 | 76 |
|  | 6 months prior | Community | 17 | 5.8 | 3 | 2 | 5 |
|  |  | Hospital | <10 | 0.2 | 12 | 3 | 47 |
|  | 12 months prior | Community | 32 | 5.8 | 6 | 4 | 8 |
|  |  | Hospital | <10 | 0.2 | 35 | 16 | 79 |
| **Bronchodilators** | Post-COVID | Community | 429 | 5.1 | 84 | 76 | 92 |
|  |  | Hospital | 36 | 0.1 | 276 | 199 | 382 |
|  | 6 months prior | Community | 292 | 5.3 | 55 | 49 | 62 |
|  |  | Hospital | 15 | 0.1 | 106 | 64 | 175 |
|  | 12 months prior | Community | 375 | 5.3 | 70 | 63 | 78 |
|  |  | Hospital | 17 | 0.1 | 118 | 73 | 190 |
| **ICS** | Post-COVID | Community | 209 | 5.3 | 40 | 35 | 46 |
|  |  | Hospital | 21 | 0.1 | 152 | 99 | 233 |
|  | 6 months prior | Community | 185 | 5.4 | 34 | 30 | 39 |
|  |  | Hospital | <10 | 0.2 | 66 | 35 | 122 |
|  | 12 months prior | Community | 199 | 5.5 | 36 | 32 | 42 |
|  |  | Hospital | <10 | 0.2 | 72 | 40 | 129 |
| **Paracetamol** | Post-COVID | Community | 119 | 5.5 | 22 | 18 | 26 |
|  |  | Hospital | 50 | 0.1 | 357 | 270 | 471 |
|  | 6 months prior | Community | 126 | 5.7 | 22 | 19 | 26 |
|  |  | Hospital | 20 | 0.2 | 129 | 83 | 200 |
|  | 12 months prior | Community | 92 | 5.7 | 16 | 13 | 20 |
|  |  | Hospital | <10 | 0.2 | 58 | 30 | 111 |
| **NSAIDs** | Post-COVID | Community | 510 | 5.1 | 100 | 92 | 109 |
|  |  | Hospital | 43 | 0.1 | 335 | 249 | 452 |
|  | 6 months prior | Community | 499 | 5.2 | 95 | 87 | 104 |
|  |  | Hospital | 23 | 0.1 | 165 | 110 | 249 |
|  | 12 months prior | Community | 577 | 5.2 | 111 | 102 | 120 |
|  |  | Hospital | 25 | 0.1 | 180 | 122 | 266 |
| **Opioids** | Post-COVID | Community | 503 | 5.1 | 99 | 90 | 108 |
|  |  | Hospital | 57 | 0.1 | 496 | 383 | 644 |
|  | 6 months prior | Community | 466 | 5.2 | 89 | 81 | 98 |
|  |  | Hospital | 43 | 0.1 | 345 | 256 | 465 |
|  | 12 months prior | Community | 521 | 5.2 | 100 | 92 | 109 |
|  |  | Hospital | 34 | 0.1 | 272 | 195 | 381 |
| **Neuropathic pain medication** | Post-COVID | Community | 270 | 5.4 | 50 | 45 | 57 |
|  |  | Hospital | 22 | 0.1 | 158 | 104 | 240 |
|  | 6 months prior | Community | 209 | 5.5 | 38 | 33 | 43 |
|  |  | Hospital | 15 | 0.2 | 99 | 59 | 164 |
|  | 12 months prior | Community | 261 | 5.5 | 47 | 42 | 53 |
|  |  | Hospital | 22 | 0.2 | 144 | 95 | 219 |

### Table S22: Health care utilization (HCU) event rates (per 100,000 person-weeks) for events identified from 2 weeks after COVID-19 diagnosis.

| **Outcome** | **Time point** | **COVID exposure group** | **Number of events** | **Time (person-weeks (PW))** | **Rate (per 100,000 PW)** | **Lower CI** | **Upper CI** |
| --- | --- | --- | --- | --- | --- | --- | --- |
| **HCU** | Post-COVID | Community | 120542 | 4.7 | 25916 | 25587 | 26250 |
|  |  | Hospital | 9943 | 0.1 | 71380 | 68707 | 74186 |
|  | 6 months prior | Community | 68499 | 5.1 | 13559 | 13337 | 13786 |
|  |  | Hospital | 5394 | 0.2 | 35586 | 33486 | 37854 |
|  | 12 months prior | Community | 94712 | 4.8 | 19813 | 19546 | 20086 |
|  |  | Hospital | 6410 | 0.2 | 42691 | 40516 | 45012 |
